# Inferring the COVID-19 infection fatality rate in the community-dwelling population: a simple Bayesian evidence synthesis of seroprevalence study data and imprecise mortality data

**DOI:** 10.1101/2021.05.12.21256975

**Authors:** Harlan Campbell, Paul Gustafson

**Affiliations:** Department of Statistics, University of British Columbia Vancouver, British Columbia, Canada

**Keywords:** COVID-19, evidence synthesis, Bayesian inference, infection fatality rate

## Abstract

Estimating the COVID-19 infection fatality rate (IFR) has proven to be particularly challenging –and rather controversial– due to the fact that both the data on deaths and the data on the number of individuals infected are subject to many different biases. We consider a Bayesian evidence synthesis approach which, while simple enough for researchers to understand and use, accounts for many important sources of uncertainty inherent in both the seroprevalence and mortality data. With the understanding that the results of one’s evidence synthesis analysis may be largely driven by which studies are included and which are excluded, we conduct two separate parallel analyses based on two lists of eligible studies obtained from two different research teams. The results from both analyses are rather similar. With the first analysis, we estimate the COVID-19 IFR to be 0.31% (95% credible interval of (0.16%, 0.53%)) for a typical community-dwelling population where 9% of the population is aged over 65 years and where the gross-domestic product at purchasing-power parity (GDP at PPP) per capita is $17.8k (the approximate worldwide average). With the second analysis, we obtain 0.32% (95% credible interval of (0.19%, 0.47%)). Our results suggest that, as one might expect, lower IFRs are associated with younger populations (and may also be associated with wealthier populations). For a typical community-dwelling population with the age and wealth of the United States we obtain IFR estimates of 0.43% and 0.41%; and with the age and wealth of the European Union, we obtain IFR estimates of 0.67% and 0.51%.

> Above all, what’s needed is humility in the face of an intricately evolving body of evidence. The pandemic could well drift or shift into something that defies our best efforts to model and characterize it.
>
> Siddhartha Mukherjee, *The New Yorker*
>
> February 22, 2021

## 1 Introduction

The infection fatality ratio (IFR), defined as the proportion of individuals infected who will go on to die as a result of their infection, is a crucial statistic for understanding SARS-CoV-2 and the ongoing COVID-19 pandemic. Estimating the COVID-19 IFR has proven to be particularly challenging –and rather controversial– due to the fact that both the data on deaths and the data on the number of individuals infected are subject to many different biases.

SARS-CoV-2 seroprevalence studies can help provide a better understanding of the true number of infections in a given population and for this reason several researchers have sought to leverage seroprevalence study data to infer the COVID-19 IFR [30]. In particular, Ioannidis [52], Levin et al. [66], Brazeau et al. [21], and O’Driscoll et al. [91] have all undertaken analyses, of varying degrees of complexity, in which they combine data from multiple seroprevalence studies with available mortality statistics to derive IFR estimates.

The analyses of both Brazeau et al. [21] and O’Driscoll et al. [91] are done using rather complex Bayesian models which rely on numerous detailed assumptions. For instance, Brazeau et al. [21] use a Bayesian “statistical age-based model that incorporates delays from onset of infection to seroconversion and onset of infection to death, differences in IFR and infection rates by age, and the uncertainty in the serosample collection time and the sensitivity and specificity of serological tests.” O’Driscoll et al. [91] employ a Bayesian ensemble model which assumes “a gamma-distributed delay between onset [of infection] and death” and assumes different risks of infection for “individuals aged 65 years and older, relative to those under 65.” While these analyses go to great lengths to account for the various sources of uncertainty in the data, the complexity of the models will no doubt make it challenging for other researchers to fit these models to different data in a constantly evolving pandemic.

In contrast, the analyses of Ioannidis [52] and Levin et al. [66] are decidedly more simple. For each seroprevalence study under consideration, Ioannidis [52] counts the cumulative number of deaths (from the beginning of the pandemic) until 7 days after the study mid-point (or until the date the study authors suggest), and divides this number of deaths by the estimated number of (previous or current) infections to obtain a study-specific IFR estimate. A “location specific” IFR estimate is then obtained by taking a weighted (by the study’s sample size) average of the study-specific IFR estimates for a given location (i.e., for a given country or state). Ioannidis [52] then calculates the median of all the location specific IFR estimates. No uncertainty interval for this estimate is provided. As such, it is impossible to determine what level of confidence one should place in Ioannidis [52]’s estimates.

The analysis of Levin et al. [66] is based on a standard frequentist random-effects meta-analysis model. For each age-group and seroprevalence study under consideration, Levin et al. [66] calculate a 95% confidence interval (CI) for a study-specific IFR by counting the cumulative number of deaths up until 4 weeks after the study mid-point and dividing this number of deaths by the estimated upper and lower bounds of the number of infected individuals. The meta-analysis model then combines each of these study-specific IFRs. While this analysis provides standard confidence intervals and is relatively straightforward, it does not take into account certain important sources of uncertainty (to be discussed in Section 2).

The analysis method we propose is simple enough for researchers to easily understand and use, while accounting for important sources of uncertainty inherent in both the seroprevalence data and the mortality data. Similar Bayesian models have been used previously for evidence synthesis of seroprevalence data for other infectious diseases (e.g., Brody-Moore [22]). We will apply the method in an analysis with the objective of estimating the average COVID-19 IFR in a community-dwelling population with a certain approximate age-composition and wealth.

A major part in any evidence synthesis is determining which studies to consider within the analysis. Determining appropriate inclusion and exclusion criteria for seroprevalence studies is a rather complicated and delicate issue when it comes to estimating the COVID-19 IFR [12, 53]. Reviewing and evaluating the merits of the hundreds of available seroprevalence studies also involves a tremendous amount of review work and time. Fortunately, both Chen et al. [29] and Arora et al. [6] have done comprehensive and thorough reviews to ascertain study quality (i.e., risk of bias). We will work from these two lists to conduct two separate parallel analyses. This approach –conducting two analyses based on two distinct and independent literature reviews– will allow us to better understand the impact of different inclusion and exclusion criteria [39]. We will review the data and how it was obtained in Section 3, following a review of the methods in Section 2. In Section 4, we summarize the results of our analysis and conclude in Section 5.

## 2 Methods

### 2.1 Bayesian model for evidence synthesis

Suppose we have data from *K* different seroprevalence studies. Then, for *k* = 1*, . . . , K*, let:

• *T_k_* be the total number of individuals tested in the *k*-th study;
• *CC_k_* be the total number of confirmed cases (of past or current infection) resulting from those tested in the *k*-th study;
• *P_k_* be the number of individuals at risk of infection in the population of interest for the *k*-th study; and
• *D_k_* be the total number of observed deaths (cumulative since pandemic onset) in the population of interest that are attributed to infection.
• We do not observe the following latent (i.e., unknown) variables; for *k* = 1*, . . . , K*, let:
• *C_k_* be the total number of infected people (cases) in the *k*-th population;
• *IR_k_* be the true infection rate (proportion of the *k*-th population which has been infected), which is the expected value of *C_k_/P_k_* ; and
• *IFR_k_* be the true underlying infection fatality rate, which is the expected value of *D_k_/C_k_* (given *C_k_* ).

We will make a series of simple binomial assumptions such that, for *k* = 1*, . . . , K*:

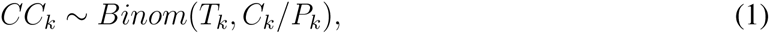

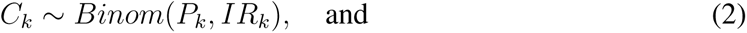

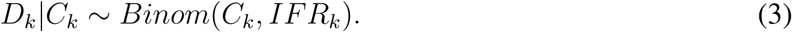

We wish to emphasize the importance of the third “*D|C*” binomial distribution above. Failing to account for the conditional distribution of the deaths given the cases may lead to inappropriately precise estimates of the IFR. For example, Streeck et al. [132] (in their original preprint (*medRxiv*, May 8, 2020)) calculate an uncertainty interval for the IFR by dividing the number of deaths (*D* = 7) by the upper and lower bounds of the 95% CI for the number of infections (95% CI for *C* = [1,551, 2,389]). Doing so, they obtain a relatively narrow 95% CI for the IFR: [0.29%, 0.45%] (= [7/1,551, 7/2,389]). In the published version of their article (*Nature Communications*, November 17, 2020), an alternative interval “accounting for uncertainty in the number of recorded deaths” is provided. This alternative interval, which essentially takes into account the *D|C* binomial distribution, is substantially wider: [0.17%; 0.77%]. In a very similar way, Levin et al. [66] also fail to take into account the *D|C* binomial distribution when estimating study-specific IFRs resulting in spuriously precise study-specific IFR estimates.

Having established simple binomial distributions for the study-specific IRs and IFRs, we define a simple random-effects model such that, for *k* = 1*, . . . , K*:

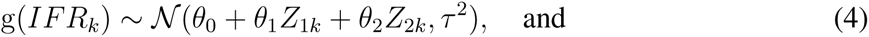

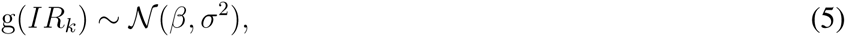

where *θ*_0_ represents the mean g(infection fatality rate), *τ* ^2^ represents between group infection fatality rate heterogeneity, *β* represents the mean g(infection rate), *σ*^2^ describes the variability in infection rates across the *K* groups, *Z*_1_*_k_* and *Z*_2_*_k_* are covariates of interest that may be related to the infection fatality rate by means of the *θ*_1_ and *θ*_2_ parameters, and g() is a given link function. In our analysis, we define g() as the complimentary log-log link function (cloglog), though there are other sensible choices including the logit and probit functions. As for the two covariates, *Z*_1_*_k_* and *Z*_2_*_k_* , we will define these as the centered and scaled logarithm of the proportion of the population aged over 65 years (65*yo_k_* ), and of the GDP (at purchasing power parity (PPP)) per capita (*GDP_k_* ), respectively.

The model is considered within a Bayesian framework requiring the specification of priors for the unknown parameters. Our strategy for priors is to assume weakly informative priors. Beta, Normal, and half-Normal priors (following the recommendations of Gelman et al. [40] and Kü mmerer et al. [63]) are set accordingly: g*^−^*^1^(*θ*) *∼ Beta*(0.3, 3); g*^−^*^1^(*β*) *∼ Beta*(1, 30); *θ*_1_ *∼ N* (0, 10); *θ*_2_ *∼ N* (0, 10); *σ ∼* half-*N* (0, 10); and *τ ∼* half-*N* (0, 10). Note that the performance of any Bayesian estimator will depend on the choice of priors and that this choice can substantially influence the posterior when few data are available [15, 64]. In the Supplementary Material 6.6, we show results obtained with an alternative set of priors as a sensitivity analysis.

### 2.2 Uncertainty in infection rates

While some seroprevalence studies report the exact number of individuals tested and the exact number of confirmed cases amongst those tested, to obtain estimates for the infection rate there are typically numerous adjustments made (e.g., adjusting for imperfect diagnostic test accuracy, adjusting for clustering of individuals within a household). For this reason, the sample size of a given study might not be a reliable indicator of its precision and weighting a study’s contribution in an evidence synthesis based solely on its sample size (as in e.g., Ioannidis [52]) may not be appropriate.

Rather than work with the raw testing numbers published in the seroprevalence studies, we calculate effective data values for *T_k_* and *CC_k_* based on a binomial distribution that corresponds to the reported 95% CI for the IR. By “inverting uncertainty intervals” in this way, we are able to properly use the adjusted numbers provided. (This is a similar approach to the strategy employed by Kümmerer et al. [63].) Tables 1 and 2 list the 95% uncertainty intervals obtained from each of the seroprevalence studies in our two parallel analyses and Tables 3 and 4 list the corresponding values for *T_k_* and *CC_k_*

**Table 1:** Seroprevalence studies selected for the analysis based on the list compiled by Chen et al. [29] (listed in alphabetical order of authors), with geographic location of sampling, sampling dates, and 95% uncertainty interval for the infection rate (IR interval). Also noted, under “In both analyses”, is whether or not each study is included in the Serotracker-based analysis (i.e., is also in Table 2). Studies with yes*^∗^* are alternate versions of studies that are included in the Serotracker-based analysis. Note that sampling for all studies took place during mid-2020, before the widespread availability of COVID-19 vaccinations.

| Authors | Location | Sampling<br>(mm/dd) | IR interval<br>(%) | In both<br>analyses |
| --- | --- | --- | --- | --- |
| Barchuk et al. [11] | Saint Petersburg, Russia | 05/27 - 06/26 | [5.60, 12.90] | yes |
| Biggs et al. [18] | DeKalb and Fulton, GA, USA (A) | 04/28 - 05/03 | [1.40, 4.50] | yes* |
| Bruckner et al. [24] | Orange County, CA, USA | 07/10 - 08/16 | [8.10, 15.50] | yes |
| Carrat et al. [27] | Ile-de-France, France | 05/04 - 06/14 | [8.90, 11.30] |  |
| Mahajan et al. [74] | Connecticut, USA | 06/10 - 07/29 | [1.70, 6.30] |  |
| Murhekar et al. (A) [86] | India | 05/11 - 06/04 | [0.34, 1.13] | yes |
| Office of National Stat [92] | England, UK (A) | 04/26 - 09/08 | [5.40, 7.10] |  |
| Petersen et al. [98] | Faroe Islands, Denmark | 04/27 - 05/01 | [0.10, 1.20] | yes |
| Pollan et al. [102] | Spain | 04/27 - 05/11 | [3.30, 6.60] | yes |
| Samore et al. [116] | Four counties in UT, USA | 05/04 - 06/30 | [0.10, 1.60] |  |
| Santos-Hovener et al. [117] | Kupferzell, Germany | 05/20 - 06/09 | [10.40, 14.00] | yes |
| Sharma et al. [120] | Delhi, India | 08/01 - 08/07 | [27.65, 29.14] | yes |
| Snoeck et al. [126] | Luxembourg | 04/15 - 05/05 | [1.23, 2.77] |  |
| Sood et al. [127] | Los Angeles County, CA, USA | 04/10 - 04/14 | [2.52, 7.07] |  |
| Statistics Jersey (A) [129] | Jersey, UK (A) | 04/29 - 05/05 | [1.80, 4.40] | yes* |
| Streeck et al. [132] | Gangelt, Germany | 03/31 - 04/06 | [12.31, 24.40] | yes |
| Stringhini et al. (A) [133] | Geneva, Switzerland (A) | 04/06 - 05/09 | [8.15, 13.95] | yes* |
| Vos et al. [141] | Netherlands | 03/31 - 05/11 | [2.10, 3.70] | yes |
| Ward et al. [145] | England, UK (B) | 06/20 - 07/13 | [5.78, 6.14] | yes |

**Table 2:**
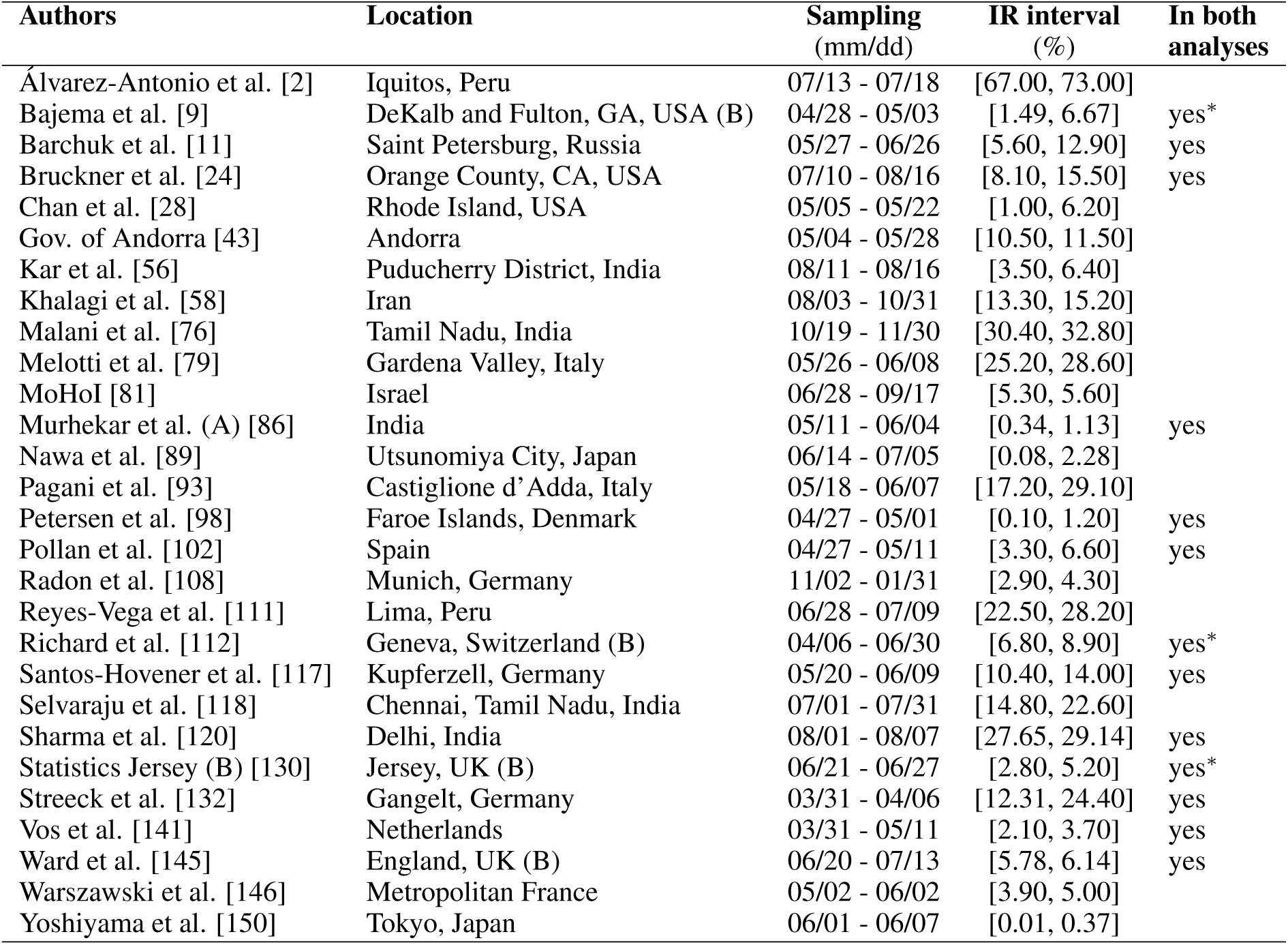
Seroprevalence studies selected for the analysis based on the list compiled by Serotracker (listed in alphabetical order of authors), with geographic location of sampling, sampling dates, and 95% uncertainty interval for the infection rate (IR interval). Also noted, under “In both analyses”, is whether or not each study is included in the Chen et al.-based analysis (i.e., is also in Table 1). Studies with yes*^∗^* are alternate versions of studies that are included in the Chen et al.-based analysis. Note that sampling for all studies took place during mid-to-late 2020, before the widespread availability of COVID-19 vaccinations.

| Authors | Location | Sampling<br>(mm/dd) | IR interval<br>(%) | In both<br>analyses |
| --- | --- | --- | --- | --- |
| Álvarez-Antonio et al. [2] | Iquitos, Peru | 07/13 - 07/18 | [67.00, 73.00] |  |
| Bajema et al. [9] | DeKalb and Fulton, GA, USA (B) | 04/28 - 05/03 | [1.49, 6.67] | yes* |
| Barchuk et al. [11] | Saint Petersburg, Russia | 05/27 - 06/26 | [5.60, 12.90] | yes |
| Bruckner et al. [24] | Orange County, CA, USA | 07/10 - 08/16 | [8.10, 15.50] | yes |
| Chan et al. [28] | Rhode Island, USA | 05/05 - 05/22 | [1.00, 6.20] |  |
| Gov. of Andorra [43] | Andorra | 05/04 - 05/28 | [10.50, 11.50] |  |
| Kar et al. [56] | Puducherry District, India | 08/11 - 08/16 | [3.50, 6.40] |  |
| Khalagi et al. [58] | Iran | 08/03 - 10/31 | [13.30, 15.20] |  |
| Malani et al. [76] | Tamil Nadu, India | 10/19 - 11/30 | [30.40, 32.80] |  |
| Melotti et al. [79] | Gardena Valley, Italy | 05/26 - 06/08 | [25.20, 28.60] |  |
| MoHoI [81] | Israel | 06/28 - 09/17 | [5.30, 5.60] |  |
| Murhekar et al. (A) [86] | India | 05/11 - 06/04 | [0.34, 1.13] | yes |
| Nawa et al. [89] | Utsunomiya City, Japan | 06/14 - 07/05 | [0.08, 2.28] |  |
| Pagani et al. [93] | Castiglione d’Adda, Italy | 05/18 - 06/07 | [17.20, 29.10] |  |
| Petersen et al. [98] | Faroe Islands, Denmark | 04/27 - 05/01 | [0.10, 1.20] | yes |
| Pollan et al. [102] | Spain | 04/27 - 05/11 | [3.30, 6.60] | yes |
| Radon et al. [108] | Munich, Germany | 11/02 - 01/31 | [2.90, 4.30] |  |
| Reyes-Vega et al. [111] | Lima, Peru | 06/28 - 07/09 | [22.50, 28.20] |  |
| Richard et al. [112] | Geneva, Switzerland (B) | 04/06 - 06/30 | [6.80, 8.90] | yes* |
| Santos-Hovener et al. [117] | Kupferzell, Germany | 05/20 - 06/09 | [10.40, 14.00] | yes |
| Selvaraju et al. [118] | Chennai, Tamil Nadu, India | 07/01 - 07/31 | [14.80, 22.60] |  |
| Sharma et al. [120] | Delhi, India | 08/01 - 08/07 | [27.65, 29.14] | yes |
| Statistics Jersey (B) [130] | Jersey, UK (B) | 06/21 - 06/27 | [2.80, 5.20] | yes* |
| Streeck et al. [132] | Gangelt, Germany | 03/31 - 04/06 | [12.31, 24.40] | yes |
| Vos et al. [141] | Netherlands | 03/31 - 05/11 | [2.10, 3.70] | yes |
| Ward et al. [145] | England, UK (B) | 06/20 - 07/13 | [5.78, 6.14] | yes |
| Warszawski et al. [146] | Metropolitan France | 05/02 - 06/02 | [3.90, 5.00] |  |
| Yoshiyama et al. [150] | Tokyo, Japan | 06/01 - 06/07 | [0.01, 0.37] |  |

**Table 3:** The Chen et al. based dataset required for the Bayesian evidence synthesis model.

| Location | $P_k$ | $D_k$<br>lower | $D_{k,}$<br>upper | $T_k$ | $CC_k$ | $65yo_k$<br>(%) | $GDP_k$<br>\$ |
| --- | --- | --- | --- | --- | --- | --- | --- |
| Saint Petersburg, Russia | 5351935 | 1596 | 5128 | 233 | 21 | 18 | 30144 |
| DeKalb and Fulton, GA, USA (A) | 1823234 | 122 | 136 | 419 | 11 | 12 | 58933 |
| Orange County, CA, USA | 3010232 | 512 | 839 | 285 | 33 | 15 | 79287 |
| Ile-de-France, France | 12213447 | 6816 | 7427 | 2414 | 243 | 14 | 82574 |
| Connecticut, USA | 2837877 | 1136 | 1179 | 251 | 9 | 22 | 80729 |
| India | 1366417750 | 4172 | 45766 | 1632 | 11 | 6 | 4735 |
| England, UK (A) | 56550000 | 25975 | 30709 | 3100 | 193 | 18 | 43310 |
| Faroe Islands, Denmark | 52154 | 0 | 0 | 592 | 3 | 17 | 60421 |
| Spain | 47351567 | 17710 | 17767 | 643 | 31 | 20 | 42362 |
| Four counties in UT, USA | 2194298 | 40 | 97 | 393 | 2 | 10 | 60050 |
| Kupferzell, Germany | 6247 | 1 | 3 | 1263 | 153 | 16 | 63885 |
| Delhi, India | 19800000 | 4188 | 26901 | 13966 | 3965 | 4 | 12817 |
| Luxembourg | 632275 | 47 | 58 | 1214 | 23 | 14 | 122166 |
| Los Angeles County, CA, USA | 10039107 | 529 | 606 | 316 | 14 | 14 | 79287 |
| Jersey, UK (A) | 107800 | 17 | 18 | 648 | 19 | 17 | 48365 |
| Gangelt, Germany | 12597 | 8 | 8 | 153 | 27 | 18 | 53751 |
| Geneva, Switzerland (A) | 504128 | 122 | 155 | 442 | 48 | 16 | 68964 |
| Netherlands | 17344874 | 2621 | 4186 | 1652 | 47 | 20 | 59685 |
| England, UK (B) | 56550000 | 29526 | 30038 | 10635 | 634 | 18 | 43310 |

**Table 4:**
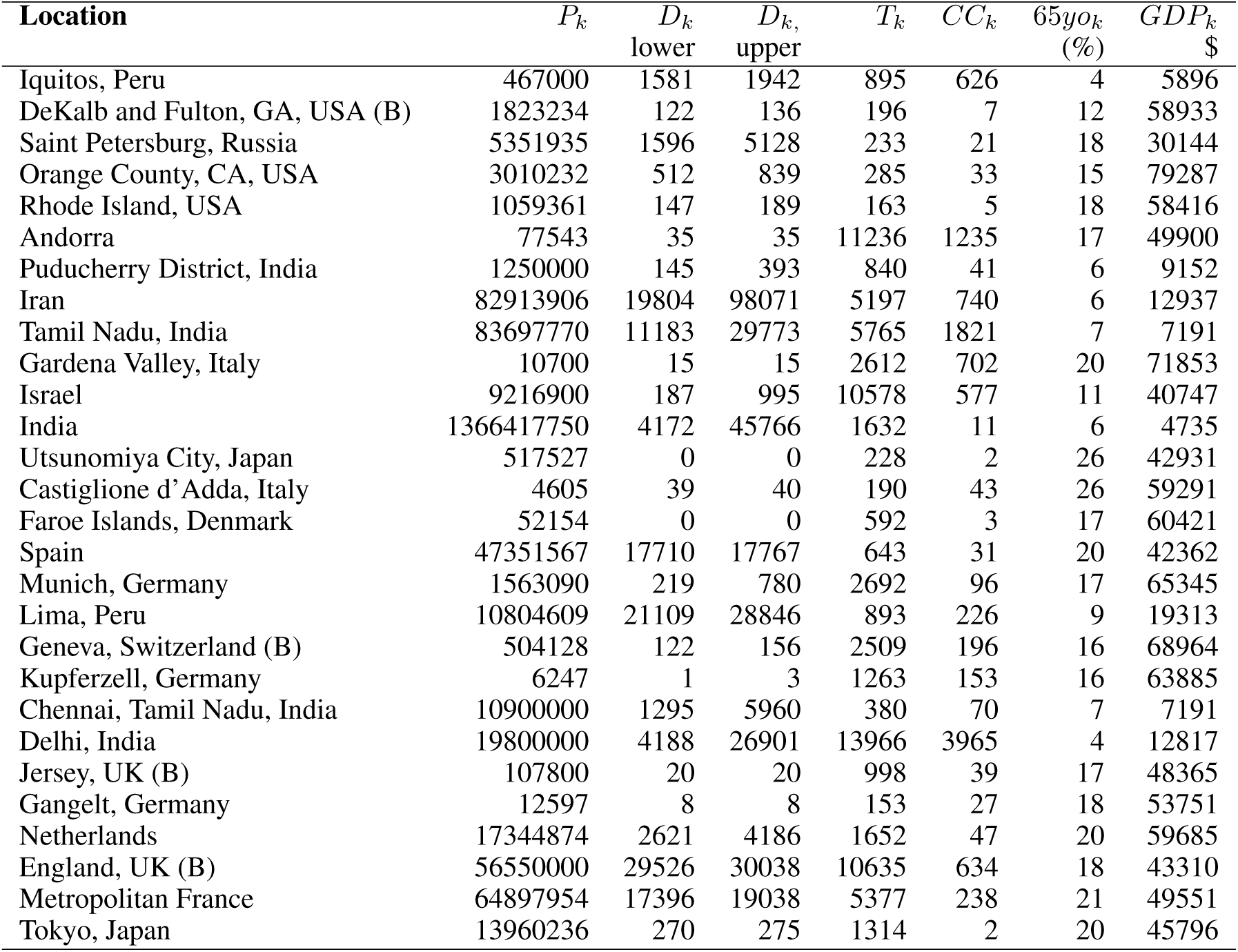
The Serotracker based dataset required for the Bayesian evidence synthesis model.

It must be noted that, as Ioannidis [52] cautions, it is possible that under our “inverting uncertainty intervals” approach, poorly conducted seroprevalence studies which fail to make proper adjustments (and thereby have spuriously narrower uncertainty intervals) receive more weight in our analysis, while high-quality studies, which make proper adjustments, are unfairly penalized. Ioannidis [52] notes that the strategy of “weighting the study-specific infection fatality rates by the sample size of each study” avoids giving more weight to studies “with seemingly narrower confidence intervals because of poor or no adjustments, while still giving more weight to larger studies.” Since we are restricting our analysis to only those supposedly high quality seroprevalence studies, we hope to largely avoid this issue. Weighting studies based on their true precision is obviously the goal in any evidence synthesis, and we recognize that this is particularly difficult when so many studies may misrepresent the precision of their estimates [19, 23].

### 2.3 Uncertainty in mortality

Matching prevalence estimates with a relevant number of fatalities is a difficult task. Prevalence estimates obtained from a seroprevalence study do not typically correspond to a specific date. Instead, these estimates will correspond to a window of time during which testing occurred. This period may be only a few days for some studies (e.g., 4 days for Petersen et al. [98]), but can also be several weeks or months for others (e.g., 135 days for Ward et al. [145]). Tables 1 and 2 list the sampling window start and end dates for each of the studies in our two parallel analyses.

Evidently, a longer sampling window will lead to greater uncertainty when it comes to establishing the relevant number of deaths. It can be difficult to account for this uncertainty and analyses will often simply select a specific date at which to count deaths based on some simple rule of thumb. For example, Ioannidis [52] considers the number of deaths at 7 days after the mid-point of the sampling window (or as the relevant number of deaths discussed by the seroprevalence study’s authors). As another example, Meyerowitz-Katz and Merone [80] take the number of deaths as recorded at 10 days after the end of the sampling window. While these two particular analytical choices are not all that different, each may lead to a substantially different number of deaths for a given study if the study was conducted during a period of time during which the number of deaths was rapidly accelerating. Levin et al. [66], who consider the number of deaths up until 4 weeks after the sampling window mid-point, acknowledge this limitation noting that: “matching prevalence estimates with subsequent fatalities is not feasible if a seroprevalence study was conducted in the midst of an accelerating outbreak.”

In order to account for the uncertainty in selecting the relevant number of deaths for a given seroprevalence study, we propose considering the number of deaths as interval censored data. Tables 3 and 4 list numbers for an interval corresponding to the number of deaths recorded 14 days after the start of the sampling window and 14 days after the end of sampling window for each seroprevalence study. While we might not know exactly what number of deaths is most appropriate, we can be fairly confident that the appropriate number lies somewhere within this interval. (Note that some intervals in Tables 3 and 4 have also been widened to account for other sources of uncertainty in the number of deaths; see details in the Supplementary Material Section 6.3). The 14 day offset allows for the known delay between the onset of infection and death, taking into consideration the delay between the onset of infection and the development of detectable antibodies [70, 149].

## 3 The data

### 3.1 Seroprevalence data

As the COVID-19 pandemic has progressed, a rapidly increasing number of SARS-CoV-2 seroprevalence studies have been conducted worldwide [6]. However, many of these studies have produced biased estimates or are otherwise unreliable due to a variety of different issues with study design, and/or with data collection, and/or with inappropriate statistical analysis [19]. We seek to restrict our analysis to high quality studies which used probability-based sampling methods. Such studies are less likely to suffer from substantial biases [123]. Based on the reviews of Chen et al. [29] and of Arora et al. [6], we compiled two separate sets of studies for analysis (these are listed in Tables 1 and 2, respectively). With the understanding that the results of an evidence synthesis may be largely driven by which studies are included/excluded, we will use these two separate sets to conduct two separate analyses. Note that the data collected for both analyses are relevant to the time before the widespread availability of COVID-19 vaccinations.

Chen et al. [29] reviewed the literature for articles published between December 1, 2019, and December 22, 2020, and identified more than 400 unique seroprevalence studies. For each of these, study quality was established using a scoring system developed on the basis of a seroepidemiological protocol from the Consortium for the Standardization of Influenza Seroepidemiology [49]. In total, Chen et al. [29] identified 38 articles which considered a sample based on a “general population” and which obtained a study quality grade of A or B (see full list in Table S8 of Chen et al. [29]). We consider these 38 articles as a starting point for inclusion for our analysis. After excluding those studies which are duplicates (n=2), those that used a “convenience” or “non-probability” based sampling method (according to the classification of Arora et al. [6]) (n=8), a study no longer considered accurate based on new information about the accuracy of the antibody test used (n=1), a study that has a very narrowly defined target population (n=1), studies for which relevant death data could not be found (n=5), and studies which did not provide a 95% uncertainty interval (n=2), we were left with a set of *K* = 19 studies for analysis; see Figure 1 and details in the Supplementary Material.

**Figure 1:**
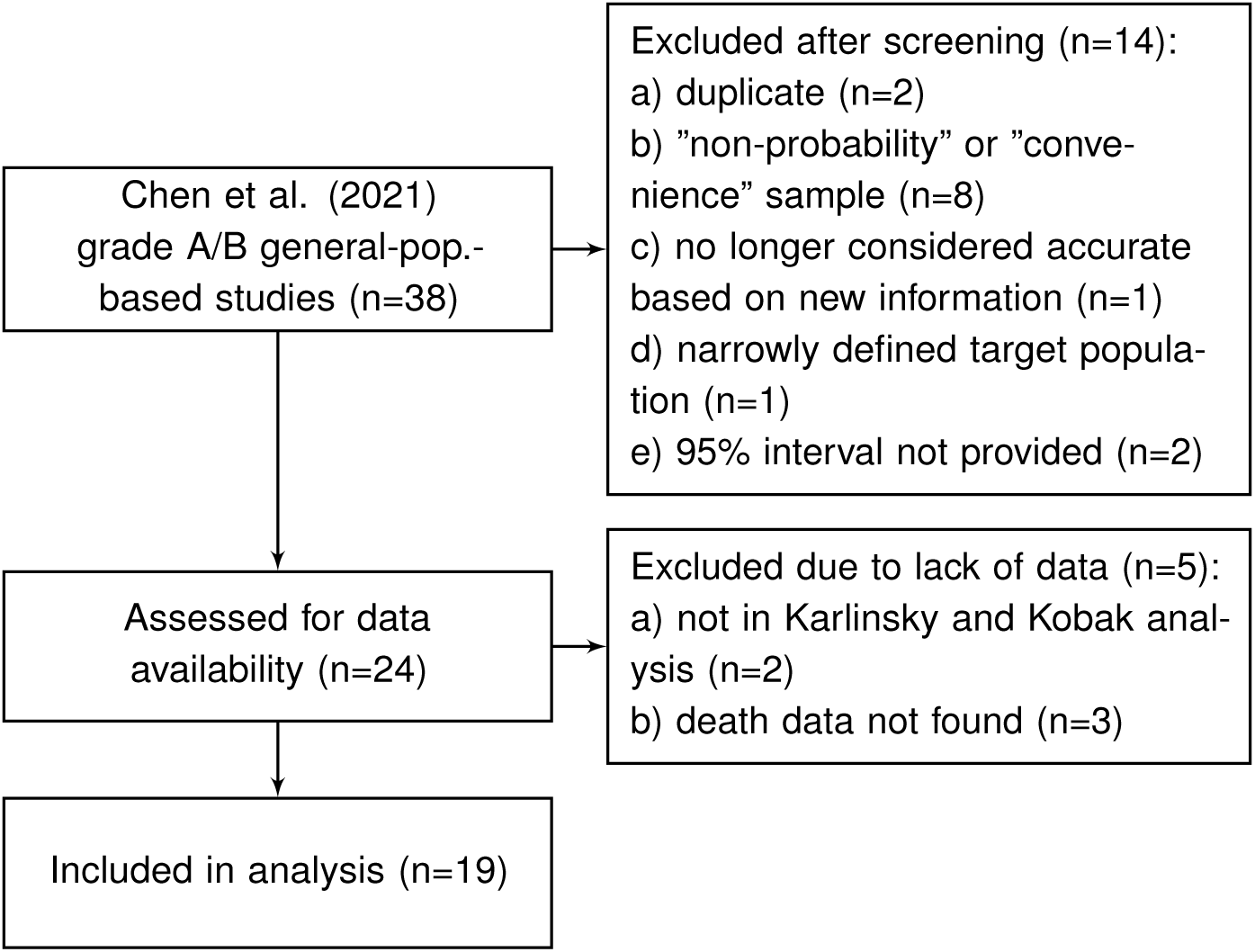
Flowchart of seroprevalence studies considered for Chen et al. -based analysis (based on the review of Chen et al. [29]).

Arora et al. [6] conducted the Serotracker “living systematic review” of COVID-19 seroprevalence studies whereby the results of the review are continuously updated on serotracker.com/data. For each study reviewed, the risk of bias was evaluated based on an assessment using the Joanna Briggs Institute Critical Appraisal Guidelines for Prevalence studies [19, 84]. For analysis, we consider the 45 studies listed on serotracker.com/data (as of June 5, 2021), that are categorized as having a “low risk of bias” and are categorized as targeting “household and community samples.” After excluding those studies which are duplicates (n=3), one study that used a “convenience” or “non-probability” based sampling method (according to the classification of Arora et al. [6]) (n=1), those studies no longer considered accurate based on new information about the accuracy of the antibody test used (n=2), those that have very narrowly defined target populations (n=2), those for which relevant death data could not be found (n=8), and those which did not provide a 95% uncertainty interval for the estimated prevalence (n=1), we are left with a set of *K* = 28 studies for analysis; see Figure 2 and details in the Supplementary Material.

**Figure 2:**
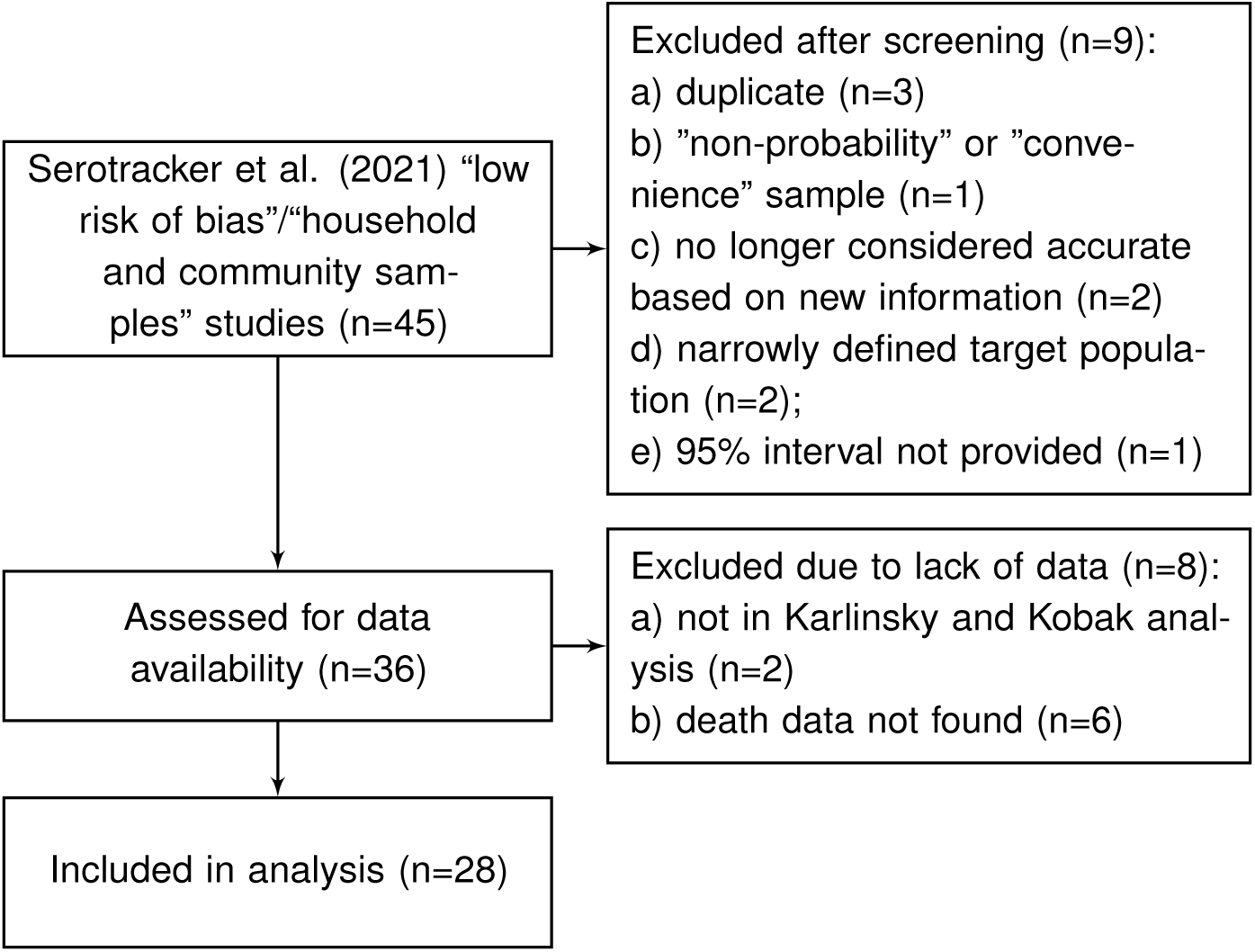
Flowchart of seroprevalence studies considered for Serotracker-based analysis (based on the Serotracker review Arora et al. [6]).

For each of the seroprevalence studies included in each of the two analysis sets, we recorded the 95% uncertainty interval for the infection rate as reported in the study article. If an article reported on multiple phases of a study (e.g., a longitudinal series of different surveys), or reported different results for different areas instead of an overall estimate (e.g., a series of different estimates for different regions), we selected only the first set of estimates. Furthermore, if a study reported more than one 95% uncertainty interval (e.g., different intervals corresponding to different adjustments and assumptions), we selected the lowest value amongst the different lower bounds and the highest value amongst the different upper bounds. These numbers are recorded in Tables 1 and 2 under *IR interval*. Based on these numbers, we calculated effective data values for the number of tests (*T_k_* ) and the number of confirmed cases (*CC_k_* ) which are listed in Tables 3 and 4 alongside population numbers (*P_k_* ) and numbers corresponding to the proportion of the population over 65 years old (65*yo_k_* ) and the GDP (PPP) per capita (*GDP_k_* ); see Supplemental Material for details and data sources.

### 3.1 Mortality data

Mortality data was obtained from various sources (e.g., academic, government, health authority); see details in Supplementary Material (Section 6.3). If a seroprevalence study referenced a specific source for mortality data, we used the referenced source for our numbers whenever possible. If no source was referenced or suggested, we considered publicly available data sources.

For many populations, there were concerns that cause of death information may be very inaccurate and lead to biased COVID-19 mortality statistics. To overcome this issue, many have suggested looking to “excess deaths” by comparing aggregate data for all-cause deaths from the time during the pandemic to the years prior [65]. For populations with a large discrepancy between the number of deaths attributed to COVID-19 and the number of excess deaths –as suggested, when possible, by a large undercount ratio (UCR) derived by Karlinsky and Kobak [57]– we used the number of deaths attributed to COVID-19 for the lower bound of the *D_k_* interval and used numbers based on excess deaths for the upper bound of the *D_k_* interval.

India, Pakistan, Palestine, Ethiopia, and China are the only countries represented in the studies that we assessed for data availability that were not included in Karlinsky and Kobak [57]’s analysis. There was evidence of substantial under-reporting of COVID-19 deaths in India [10, 104] while little could be gathered about the reliability of official mortality data for Pakistan, Palestine^1^, Ethiopia^2^, and China (but do see [72] and [137]). As such, we excluded the Qutob et al. [107] (“Palestinian population residing in the West Bank”) and He et al. [47] (“Wuhan, China”) studies from the Serotracker-based analysis and excluded the Alemu et al. [1] (“Addis Ababa, Ethiopia”) and the Nisar et al. [90] (“Two neighborhoods of Karachi, Pakistan”) studies from the Chen et al.-based analysis. For India, Mukherjee et al. [83] and Purkayastha et al. [105] estimate UCRs for the entire country as well as for each individual Indian state and union territory. We used these UCRs to adjust the death numbers for seroprevalence studies in India (see Supplementary Material for details).

There are two countries represented within our data that were identified by Karlinsky and Kobak [57] as having large discrepancies between the official number of deaths attributed to COVID-19 and the number of excess deaths Iran (with UCR=2.4) and Russia (with UCR=4.5). As such, for the Barchuk et al. [11] (“Saint Petersburg, Russia”) and for the Khalagi et al. [58] (“Iran”), we used numbers based on excess deaths (see Supplementary Material for details).

Finally, our target of inference is the IFR for the community-dwelling population and does not apply to people living in long-term care (LTC) facilities (also known as “nursing homes” or, in France as “Établissement d’hébergement pour personnes âgées dépendantes” (EHPAD)). The spread of COVID-19 is substantially different in LTC facilities than in the general population, and residents of LTC facilities are particularly vulnerable to severe illness and death from infection; see Danis et al. [31]. With this in mind, we made adjustments (when appropriate/possible) to the mortality numbers used in our analysis in order to exclude deaths of LTC residents; see Supplementary Material for details. Modeling the spread and mortality of COVID-19 within LTC facilities will require unique approaches and should be considered in a separate analysis; see the recommendations of Pillemer et al. [100].

## 4 Results

The model as described in Section 2, was fit to the two datasets as described in Section 3. We fit the model using JAGS (just another Gibbs sampler) [62], with 5 independent chains, each with 2 million draws (20% burn-in, thinning of 100); see Supplementary Material (Section 6.4) for details and JAGS code.

We report posterior median estimates and 95% highest probability density (HPD) credible intervals (CrI). Figure 3 (for the Chen et al.-based analysis) and Figure 4 (for the Serotracker-based analysis) plot the point estimates and credible intervals obtained for *IFR_k_* , for *k* in 1*, . . . , K*, respectively; see Figures 5 and 6 for *IR_k_* are in the Supplementary Material. In these figures the seroprevalence studies are listed in order of their “fitted” IFR values (the posterior median of *g^−^*^1^(*θ*_0_+*θ*_1_*Z*_1_*_k_* +*θ*_2_*Z*_2_*_k_* ), for *k* in 1*, …, K*, marked on the plot by the *×* symbols). Results obtained for the other model parameters are listed in Table 5.

**Figure 3:**
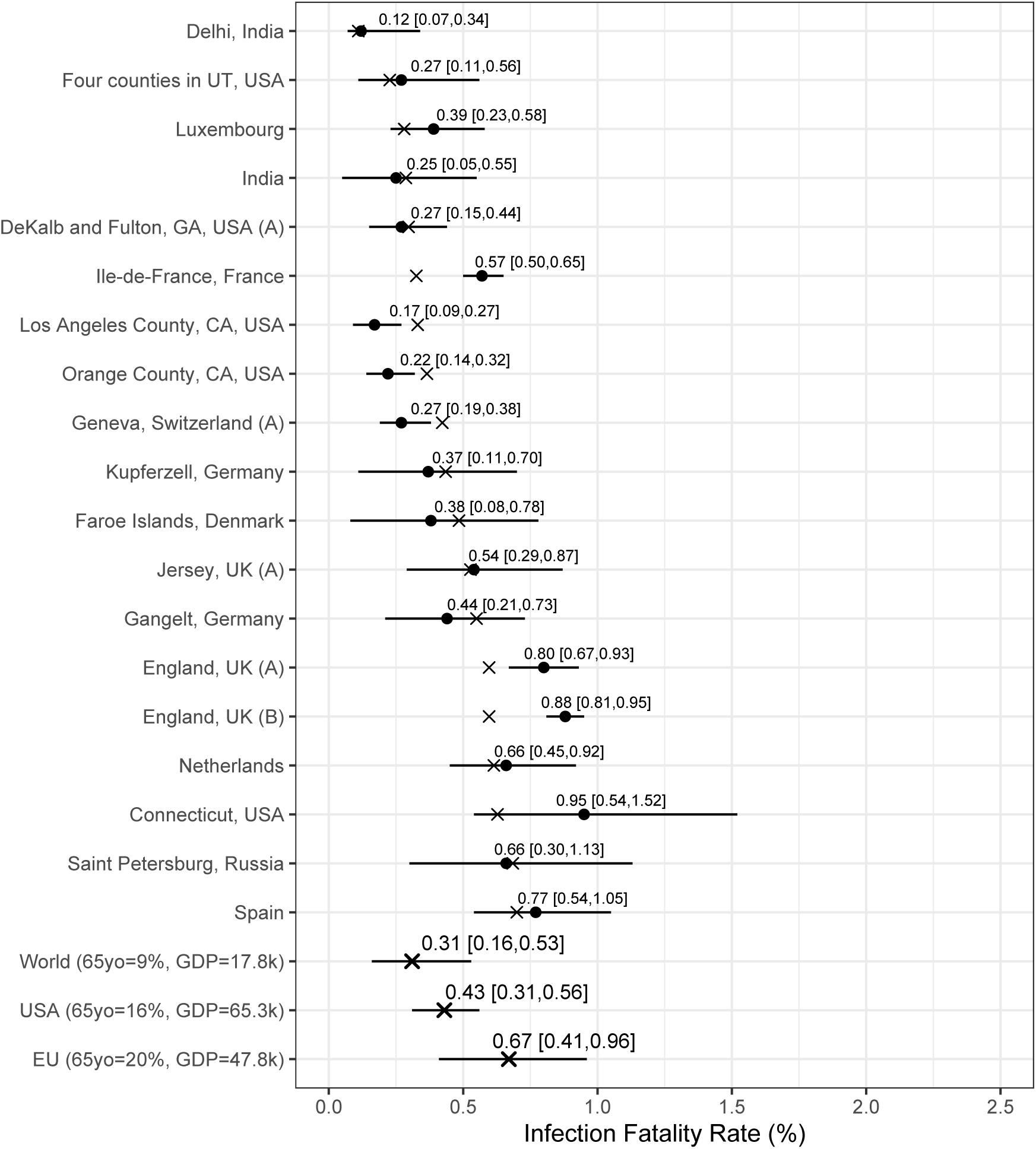
Results from the Chen et al.-based analysis: posterior median estimates (black circles) for the *IFR_k_* variables (for *k* = 1*, . . . ,* 19) with 95% HPD CrIs. Studies are listed from top to bottom in order of increasing fitted values (these values are indicated by ). Also plotted, under the labels “World (65 yo=9%, GDP=17.8k)”, “USA (65 yo=16%, GDP=65.3k)”, “EU (65 yo=20%, GDP=47.8k)”, are the posterior median estimate and 95% HPD CrIs for the typical IFR corresponding to values for the proportion of the population aged 65 years and older of 9% and for GDP per capita of $17,811 (the worldwide values), of 16% and of $65,298 (the USA values), and of 20% and of $47,828 (the EU values).

**Figure 4:**
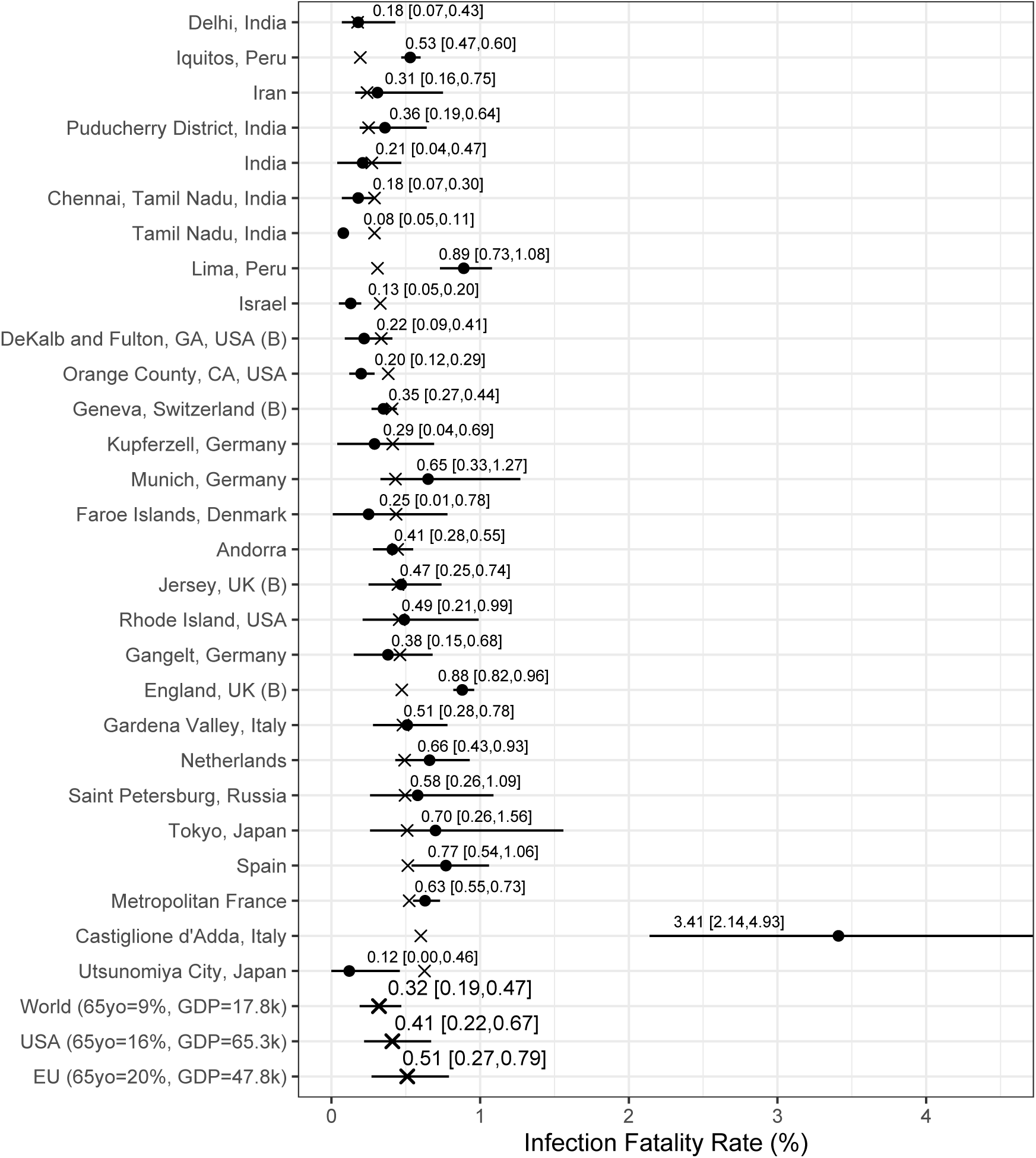
Results from the Serotracker-based analysis: posterior median estimates (black circles) for the *IFR_k_* variables (for *k* = 1*, . . . ,* 28) with 95% HPD CrIs. Studies are listed from the top to the bottom in order of increasing fitted values (these values are indicated by ). Also plotted, under the labels “World (65 yo=9%, GDP=17.8k)”, “USA (65 yo=16%, GDP=65.3k)”, “EU (65 yo=20%, GDP=47.8k)”, are the posterior median estimate and 95% HPD CrIs for the typical IFR corresponding to values for the proportion of the population aged 65 years and older of 9% and for GDP per capita of $17,811 (the worldwide values), of 16% and of $65,298 (the USA values), and of 20% and of $47,828 (the EU values).

**Table 5:**
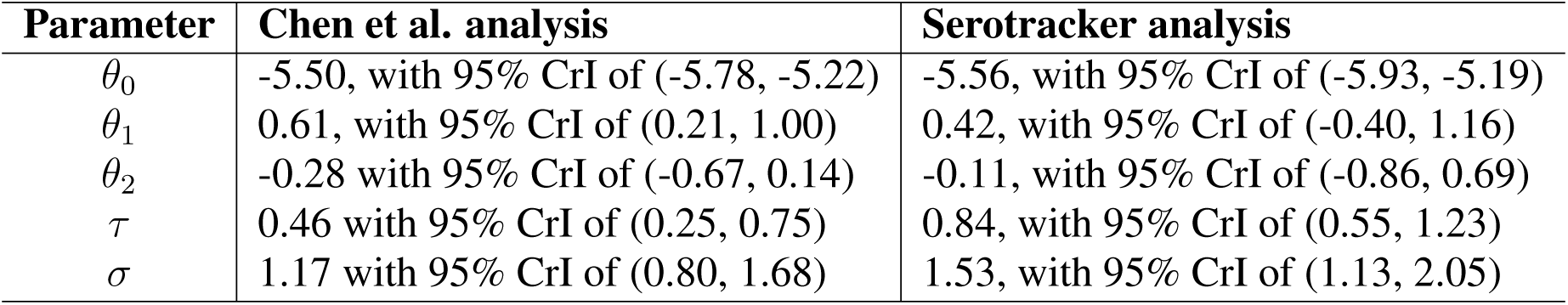
Parameter estimates (posterior medians and 95% HPD credible intervals) obtained from the Chen et al.-based analysis and the Serotracker-based analysis.

In general, the Chen et al.-based analysis and the Serotracker-based analysis provide mostly similar results. Notably, the Serotracker-based analysis considers a much more geographically diverse set of seroprevalence studies and several studies that appear to be prominent outliers (e.g., “Tamil Nadu”, “Castiglione d’Adda, Italy”, and “Utsunomiya City, Japan”), see Figure 4. These outliers could be due to infection rates in these populations being markedly different for the elderly relative to the general population.^3^ With regards to heterogeneity, fitting the model without any covariates, one obtains 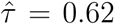 (Chen et al.-based analysis) and 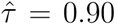 (Serotracker-based analysis). This suggests that the two covariates, 65*yo_k_* and *GDP_k_* , account for approximately 45% (= (0.62^2^ *−* 0.46^2^)*/*(0.62^2^); Chen et al.-based analysis) and 13% (= (0.90^2^ *−* 0.84^2^)*/*(0.90^2^); Serotracker-based analysis) of the heterogeneity in the IFR.^4^

Our estimates of 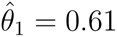 (Chen et al.-based analysis) and 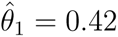 (Serotracker-based analysis) suggest that older populations are more likely to have higher IFRs. This is as expected since age is known to be a very important risk factor [148, 152]. Our estimate of 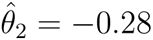 (Chen et al.-based analysis) and 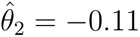 (Serotracker-based analysis) suggest that wealthier populations may be more likely to have lower IFRs. However, the wide credible intervals obtained for the *θ*_2_ parameter (in both analyses) suggest a much less definitive conclusion. There are several reasons which might explain this result. As with any observational data analysis, the estimate of *θ*_2_ may suffer from bias due to unobserved confounding and statistical power may be compromised by the presence of outliers and insufficient heterogeneity in the GDP per capita metric across the different populations included in our analyses.

We can infer (by determining the posterior median of *g^−^*^1^(*θ*_0_ + *θ*_1_*z*_1_*_∗_* + *θ*_2_*z*_2_*_∗_*), for selected values of *z*_1_*_∗_* and *z*_2_*_∗_*) the *typical* IFR amongst populations (be they included in our study or not) having a given proportion of the populace aged over 65 and a given GDP per capita. Thus we calculate posterior point and interval estimates corresponding to age and wealth values that match the population of the entire world (World), the United States (USA), and the European Union (EU) (as listed by the World Bank’s World Development Indicators (WDI)); see “World”, “USA”, and “EU” rows in Figures 3 and 4. For 65yo = 9% and GDP = $17,811, the approximate worldwide values, we obtain, from the Chen et al.-based analysis, an across-population average IFR estimate of 0.31%, with a 95% CrI of (0.16%, 0.53%). With the Serotracker-based analysis, we obtain a similar estimate of 0.32%, with a 95% CrI of (0.19%, 0.47%). For 65yo = 16% and GDP = $65,298, the USA values, we obtain across-population average IFR estimates of 0.43%, with a 95% CrI of (0.31%, 0.56%) (Chen et al.-based analysis) and of 0.41%, with a 95% CrI of (0.22%, 0.67%) (Serotracker-based analysis). Finally, for 65yo = 20% and GDP = $47,828, the EU values, we obtain across-population average IFR estimates 0.67%, with a 95% CrI of (0.41%, 0.96%) (Chen et al.-based analysis) and of 0.51%, with a 95% CrI of (0.27%, 0.79%) (Serotracker-based analysis). Note that for the “World” predictions, the Serotracker-based analysis has the more precise estimates, while the Chen et al.-based estimates are more precise for the “USA” predictions.

While the infection-rate estimates obtained from the seroprevalence studies should be relatively reliable (due to having satisfied the risk of bias assessments of either Chen et al. [29] or Arora et al. [6]), the mortality data we collected may be less reliable depending on the target population and source. The data which were not obtained from official and reliable sources may be particularly suspect. With this in mind, as a sensitivity analysis, we repeated both analyses with these data excluded; see results in Figures 7 and 8 in the Supplementary Material Section 6.6. Without the excluded studies, we are unable to provide a reasonable “World” estimate (see the extremely wide credible intervals). However, the “USA” and “EU” estimates are relatively similar. We also repeated the two analyses using a different set of priors to verify that our results were not overly sensitive to our particular choice of priors. The results of this alternative analysis are very similar to the results of our original analyses; see Figures 9 and 10 in the Supplementary Material Section 6.6.

Our estimates are somewhat similar to those obtained in other analyses. Brazeau et al. [21], using data from 10 representative seroprevalence studies (identified after screening 175 studies), infer “the overall IFR in a typical low-income country, with a population structure skewed towards younger individuals, to be 0.23% (0.14%-0.42% 95% prediction interval range).” For a “typical high income country, with a greater concentration of elderly individuals,” Brazeau et al. [21] obtain an estimate of 1.15% (95% prediction interval of 0.78%-1.79%). Ioannidis [52], using data from seroprevalence studies with sample sizes greater than 500 (and including deaths of LTC residents), obtains a “median infection fatality rate across all 51 locations” of 0.27% and (and of 0.23% following an adhoc correction to take into account “that only one or two types of antibodies” may have been tested in some seroprevalence studies). Levin et al. [66], who restricted their analysis to populations in “advanced economies” (and included deaths of LTC residents) provide age-group specific estimates and country-specific estimates. For instance, for the 45–54 year old age group, Levin et al. [66] estimate the IFR to be 0.23% (95% CI of 0.20%–0.26%), and for the 55–64 year old age group, 0.75% (95% CI of 0.66%–0.87%). For Spain, Levin et al. [66] estimate an IFR of 1.90%. For comparison, we estimate the IFR for the community-dwelling population (i.e., excluding deaths of LTC residents) of Spain to be 0.77% (in both analyses). This is similar to the 0.83% estimate obtained by Pastor-Barriuso et al. [94] and the 0.75% estimate obtained by Brazeau et al. [21] (both of these excluding deaths of LTC residents).

Specifically with regards to the United States, Sullivan et al. [135] estimate the IFR for adults to be 0.85% (95% CrI of 0.76%-0.97%) based on a US nationwide seroprevalence survey conducted between August and December, 2020.^5^ Pei et al. [95] using a rather complex Bayesian “metapopulation” model conclude that, for the United States during 2020, the IFR likely “decreased from around 1% in March to about 0.25% in December.” For comparison, our “USA” predictions of 0.43% and of 0.41% are based on data obtained mostly between April, 2020 and August, 2020 (see dates in Tables 1 and 2).

## 5 Conclusion

Estimation of the IFR can be incredibly challenging due to the fact that it is a ratio of numbers where both the numerator and the denominator are subject to a wide range of biases. Our proposed method seeks to address some of these biases in a straightforward manner. In our analysis, proper handling of the various sources of uncertainty was a primary focus [151].

With regards to the numerator, we considered the number of deaths as interval censored data so as to account for the uncertainty in selecting the most relevant number of deaths. While we consider this an improvement over other methods that use a single fixed number, we acknowledge that the specific choice of a 14 day offset is somewhat arbitrary and that the data for deaths also suffer from other sources of bias. Ioannidis [53] notes that the time between infection and death may vary substantially “and may be shorter in developing countries where fewer people are long-sustained by medical support.” In addition to official numbers, we used mortality data based on “excess deaths” statistics for Russia and Iran, since official mortality statistics appeared to be potentially highly inaccurate. We also used adjusted mortality numbers for India based on the best available information. These adjustments are certainly not perfect and we note that “excess deaths” statistics may also suffer from substantial inaccuracies [82].

With regards to the denominator, we looked to data from “high-quality” seroprevalence studies in an effort to avoid biased estimates. However, these data are also not perfect. Seroprevalence studies are severely limited by the representativeness of the individuals they test. Certain groups of individuals who may have very high infection rates are unlikely to be tested in a seroprevalence study (e.g., homeless people). On the other hand, those individuals who have reason to believe they may have been infected, may be more likely to volunteer to participate in a seroprevalence study [123]. It is also likely that seroreversion (loss of detectable antibodies over time) may lead to a seroprevalence study underestimating the true number of infections if the time between the main outbreak and the subsequent antibody testing is substantial [96]. Notably, Axfors and Ioannidis [7] employ a “X-fold”-based correction factor to adjust seroprevalence estimates for this type of bias.

The need to improve the quality and reporting of seroprevalence studies cannot be overemphasized.^6^ A major limitation of evidence synthesis is often summarized by the expression “garbage in, garbage out” [34], meaning that if one includes biased studies in one’s analysis, the analysis results will themselves be biased [122]. In our two analyses, we only included data from 19 and 28 out of potentially hundreds of seroprevalence studies due primarily to the fact that so few studies were considered reliable and at low risk of bias. Excluding low-quality/biased studies from our analysis was necessary, at least to a certain degree, in order to obtain valid estimates. However, as a consequence of our strict exclusion criteria, much of the world’s population is severely under-represented in our data. In related work, Levin et al. [67] review the available literature and “informally assess studies for risk of bias” in an attempt to estimate the COVID-19 IFR specifically for developing countries. If the quality of studies were to be correlated with unmeasured factors that impact the IFR, excluding studies based on their perceived quality could lead to unmeasured confounding at a meta-analytic level [54]. Novel methods which allow evidence syntheses to appropriately incorporate biased data are urgently needed. Recently, Campbell et al. [25] proposed a partially identified model to combine seroprevalence study data with biased data from official statistics.

Outside of biased data, perhaps the foremost challenge in evidence synthesis using observational data is that necessarily one is forced to make an array of design choices around inclusion/exclusion criteria, statistical modeling approaches, and prior specifications [39]. With the two separate analyses and the various additional sensitivity analyses, we were quite encouraged by the stability of our results to perturbations of these inputs.

Reducing the uncertainty around the severity of COVID-19 was of great importance to policy makers and the public during the early stages of the pandemic [35, 51, 71] and immense efforts have been made in the collection and analysis of data (e.g., Williamson et al. [148]). And yet, even after more than a year, there is still a large amount of uncertainty and unexplained heterogeneity surrounding the COVID-19 IFR, particularly with respect to populations in less affluent countries. While a certain amount of heterogeneity is to be expected [48], identifying factors associated with higher IFRs is the ultimate goal and investigating potential variables that can account for the observed heterogeneity may lead to important insights [16, 54].

We prioritized simplicity in our modeling so as to promote transparency in our findings, and to facilitate adaptations to similar, but not identical, data structures. While “simple” is a relative term, note that the entire dataset used for our analyses fits on a single page (in Tables 3 and 4) and that the entire JAGS MCMC code fits on less than a single page (see Supplementary Material). One model extension that could be pursued would involve age stratification of IFR.

Including age-stratification in the model could represent a substantial improvement given that infection in some populations is far from homogeneous (e.g., about 95% of Singapore’s COVID-19 infections were among young migrant workers (as of September, 2020), which explains the incredibly low case fatality rate [37]). If a factor, such as age, impacts both the risk of infection and the risk of death given infection, then estimating the IFR as we have done in our analysis could be subject to confounding [147]. Age-group specific seroprevalence/mortality data is available for certain geographic areas [113] (although not always consistently reported) and such data could inform an extended version of our model, thereby offering an alternative to the approach described by Levin et al. [66] for estimating age-group specific IFRs.

Finally, we must emphasize that the IFR is a moving target. As the pandemic changes, so to does the IFR. Our estimates are based on data from 2020, most of which were obtained more than a year ago. It is likely that, with continual viral mutation of SARS-CoV-2, advances in treatment, and the availability of vaccines, the current IFR in many places is now markedly different than it was earlier in 2020 [95, 99, 143].

**Key Messages**

• The COVID-19 IFR is estimated to be about 0.32% for a typical community-dwelling population where 9% of individuals are over 65 years old and where the GDP per capita is $17.8k (the approximate worldwide averages). For a typical community-dwelling population with the age and wealth of the United States we estimate the IFR to be approximately 0.42%.
• Any estimation of the COVID-19 IFR should take into account the various uncertainties and potential biases in both the mortality data and the seroprevalence data.
• Bayesian methods with interval censoring are well suited for complex evidence synthesis tasks such as estimating the COVID-19 IFR.

## Availability of data and material

Data used for the analysis is available in the Supplementary Material and at https://github.com/harlanhappydog/BayesianSeroMetaAnalysis.

## Code availability

Code used for the analysis is available in the Supplementary Material and at https://github.com/harlanhappydog/BayesianSeroMetaAnalysis.

## Data Availability

The data collected for the analysis is available at: https://tinyurl.com/awskkwkn

## Acknowledgments

This work was supported by the European Union’s Horizon 2020 research and innovation programme under ReCoDID grant agreement No 825746 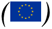 *and by the Canadian Institutes of Health Research, Institute of Genetics (CIHR-IG) under Grant Agreement No 01886-000. We wish to thank Andrew Levin, Thomas Debray, Valentijn de Jong, and Thomas Jaenisch for their valuable feedback. We also wish to thank Ariel Karlinsky, Nana Owusu-Boaitey, Lauren Maxwell, and Sayali Arvind Chavan for help with data collection. Finally, thank you to the many fellow researchers who helped by providing information about various data including, amongst others, Maurizio Napolitano, Shiwani Mahajan, Miguel Quartin, Stefano Lombardo, and Timon Gaertner*.

## 6 Supplementary Material

### 6.1 Excluded studies - Chen et al.-based analysis

Figure 1 shows a flowchart of the literature search and Table 6 lists all the excluded studies. Among the 38 articles obtained from the Chen et al. review, four studies represented results from different phases of the same study. For each of these we considered only the data from the earliest phase of the study. Stringhini et al. [133] and Richard et al. [112] are two publications that report the earlier and later phases, respectively, of the same study of Geneva, Switzerland. We considered only data from the earlier phase as reported in Stringhini et al. [133]. Murhekar et al. [86] and Murhekar et al. [87] are two publications that report the earlier and later phases, respectively, of the same study in India. We considered only data from the first phase as reported in [86].

Note that, while similar in many ways, Ward et al. [145] and Office of National Statistics [92] are two different large-sample studies. The Ward et al. [145] study is based on the “REACT-2” survey which is led by Imperial College London, while the Office of National Statistics [92] study is based on the “Coronavirus (COVID-19) Infection Survey” which is conducted by a partnership between the University of Oxford, University of Manchester, Public Health England and Wellcome Trust. Spiers [128] discusses the differences between the two surveys.

We excluded eight studies that used “non-probability” or “convenience” sampling methods (e.g, studies in which participants were recruited using social media, or recruited in shopping centers). Specifically, we excluded: (1) McLaughlin et al. [78] who sampled individuals from a list of volunteers (Arora et al. [6] classifies the sampling method for this study as “self-referral”); (2) Rosenberg et al. [114] who sampled individuals at grocery stores (“convenience” [6]); (3) Appa et al. [5] who recruited volunteers with support from Bolinas community leaders (“self-referral” [6]); (4) Bendavid et al. [14] who recruited participants by placing targeted advertisements on Facebook (“stratified non-probability” [6]); (5) Gudbjartsson et al. [44] (“convenience” [6]); (6) Borges et al. [20] (“convenience” [6]); (7) Ling et al. [69] (“self-referral” [6]); and (8) Naranbhai et al. [88] (“convenience” [6]). Due to new information about the sensitivity of the Wondfo antibody tests (Silveira et al. [125]: “Our findings cast serious doubts about the use of this brand of rapid tests for epidemiological studies.”), we excluded the Hallal et al. [45] Brazil study (but do see: Marra and Quartin [77]). One study (Malani et al. [76]) was excluded due to a narrowly defined target population.

Two additional studies were not included because the articles failed to report 95% uncertainty intervals for the estimated infection rate in the target population [75, 144] and five additional studies were not included due to unavailable reliable mortality data for the specific target populations [1, 90, 103, 136, 119].

**Table 6:**
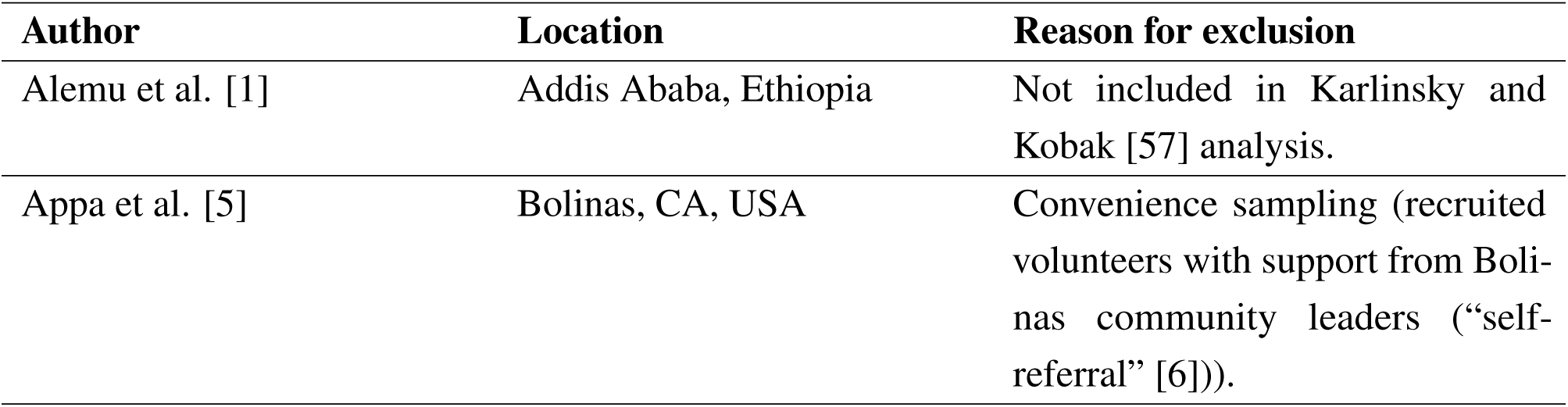
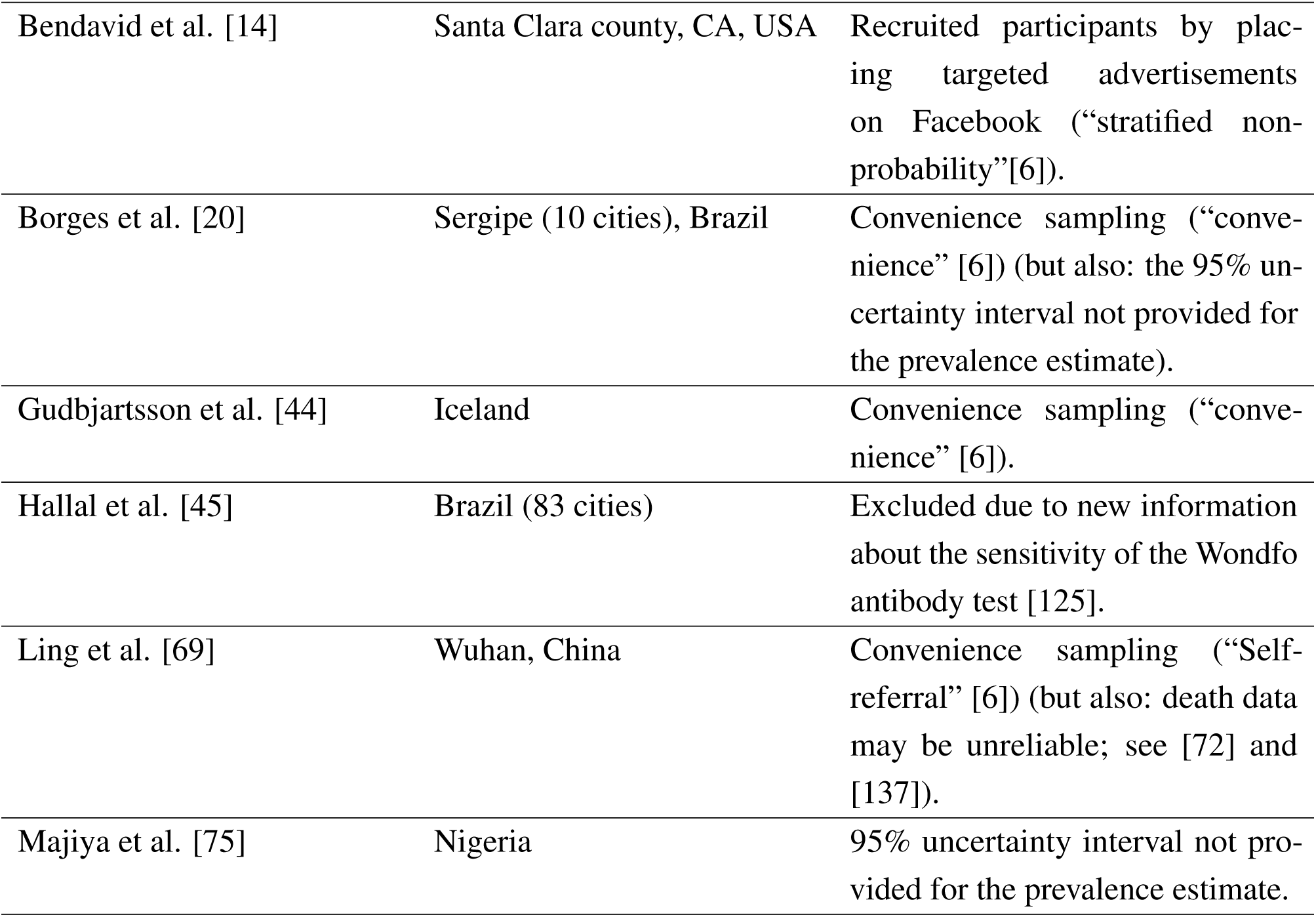
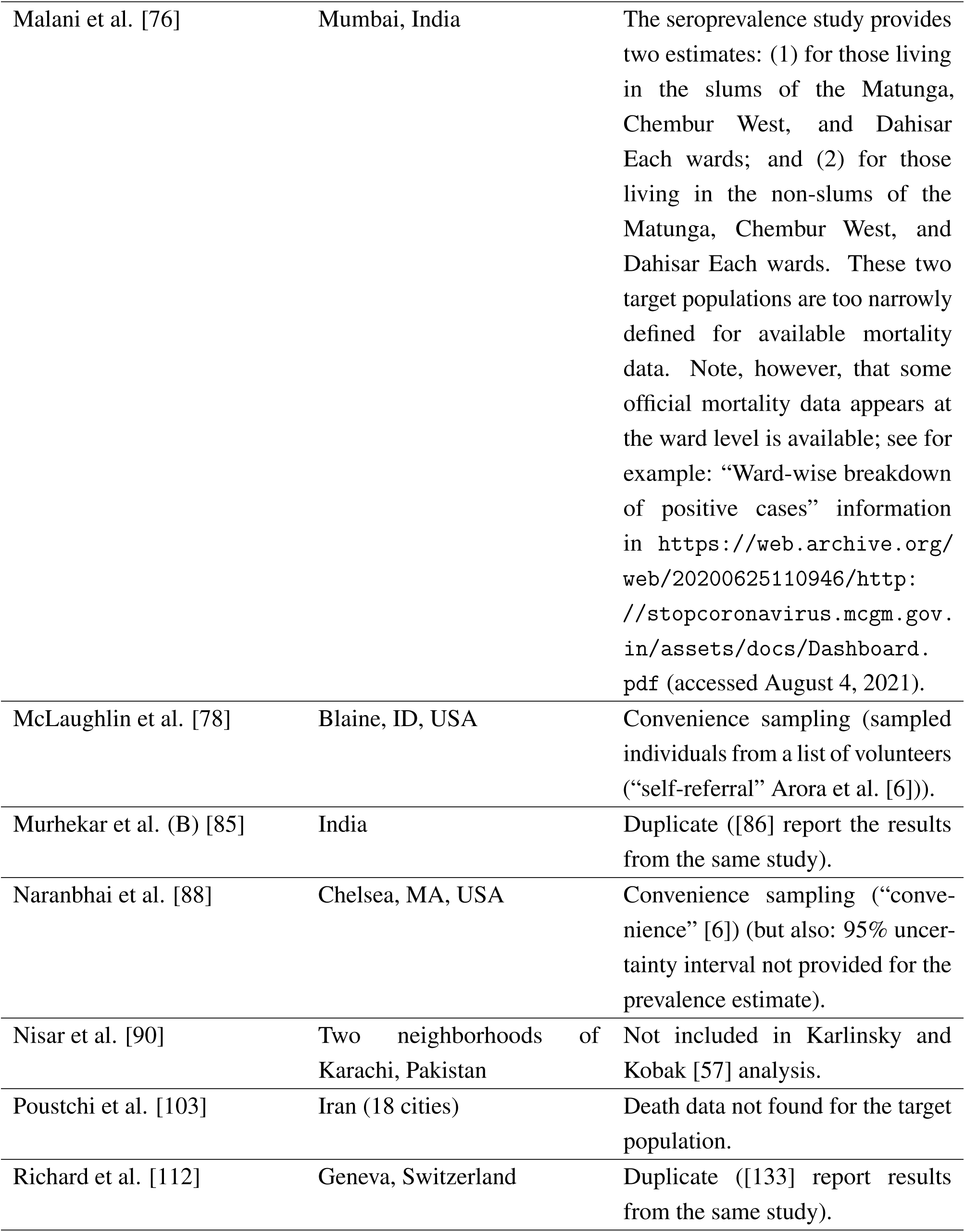
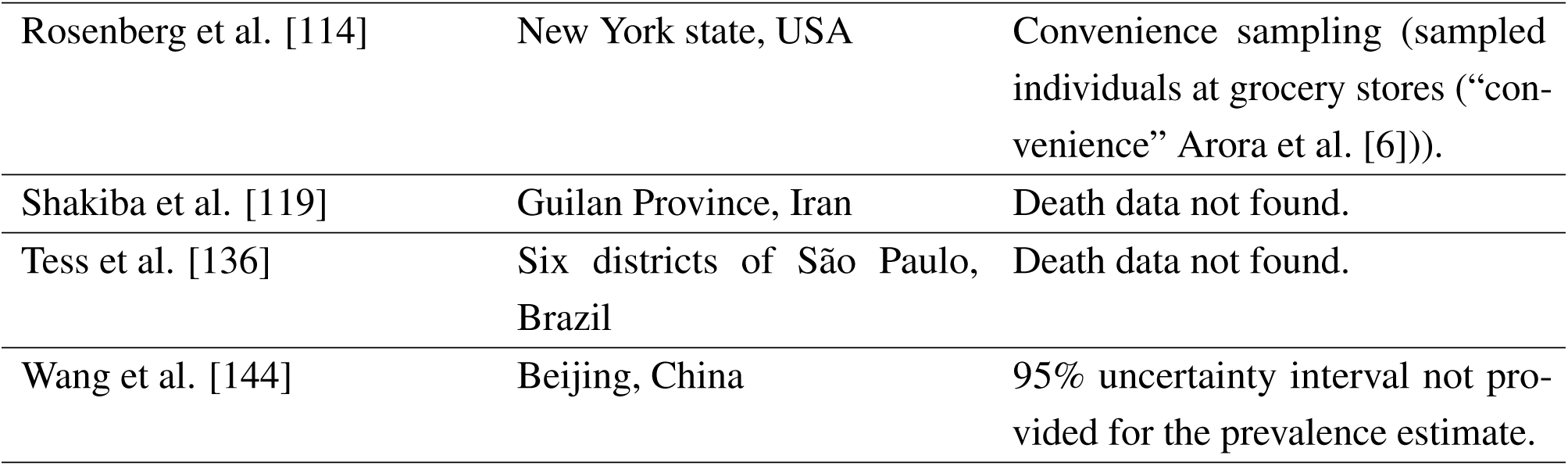
List of excluded studies and reason for exclusion for the Chen et al. - based analysis.

### 6.2 Excluded studies - Serotracker-based analysis

Figure 2 shows a flowchart of the literature search and Table 7 lists all the excluded studies. Among the 45 articles obtained from the Serotracker review, six studies represented results from different phases of the same study. For each of these we considered only the data from the earliest phase of the study. For instance, Murhekar et al. [86] and Murhekar et al. [87] report the results from an earlier and a later phase of the same study in India. We only include Murhekar et al. [86] for our analysis. We excluded one additional study that used “convenience” sampling methods [131] and two studies that used the Wondfo antibody test [68, 124] (without adequate adjustment given the recent information about the sensitivity of the Wondfo test Silveira et al. [125]). We excluded two additional studies that had a narrowly defined target population[8, 139].

One additional study was not included because the article failed to report a 95% uncertainty interval for the estimated infection rate in the target population (Truc and Gervino [138] focused on estimating the longitudinal changes in antibodies levels rather than the prevalence rate in Cogne, Italy.). Note that the “Andorra” study [43] also failed to report a 95% uncertainty interval for the estimated infection rate in the target population. However, we chose to exceptionally include the study due to the fact that it represents one of the largest seroprevalence studies conducted: a total of 70,494 inhabitants (90.9% of the population of Andorra) participated in the study. In their published article Royo-Cebrecos et al. [115] explain that 95% confidence intervals are not provided because such intervals “would be extremely narrow and potentially misleading given that they do not account for the potential bias that non-participating individuals could cause on our central seroprevalence estimate.” We therefore defined, in order to be very cautious, a very wide (yet entirely arbitrary) 95% confidence interval of [10.5%, 11.5%] around the 11.0% point estimate. Eight additional studies were not included due to unavailable reliable mortality data for the specific target populations.

**Table 7:** List of excluded studies and reason for exclusion for the Serotracker-based analysis.

| **Author** | **Location** | **Reason for exclusion** |
| --- | --- | --- |
| Backhaus et al. [8] | Dusseldorf, Germany | Sample not representative of overall population: only "young people between the ages of 18-30" and individuals in the "fire brigade and rescue services". |
| Beaumont et al. [13] | Three neighbourhoods in the city of Perpignan, France | Death data not found for the target population (three specific neighbourhoods (Saint-Jacques, Haut-Vernet, and Nouveau Logis) in the city of Perpignan, France). |
| Gégout-Petit et al. [38] | Grand Nancy metropolitan area, France | Death data not found; however, note that coronadatascraper.com does have data for the larger region of Meurthe-et-Moselle, Grand Est, France. |
| He et al. [47] | Wuhan, China | Not included in Karlinsky and Kobak [57]'s analysis and see [72] and [137] who discuss issues as to why this is particularly challenging and controversial data to validate. |
| Huamaní et al. [50] | Cusco, Peru | Death data not found for the target population ("three settings in Cusco: (1) Cusco city, (2) the periphery of Cusco, and (3) Quillabamba city"). |
| Khan et al. [60] | Vale Kashmir, India | Death data not found (official death numbers can be found on "Official Twitter handle of Department of Information and Public Relations, Govt of Jammu & Kashmir ("@diprjk"). While not specifically referenced, the numbers reported by this source are consistent with those used in Khan et al. [59].) Mukherjee et al. [83] does not provide an estimated underreporting factor for Kashmir noting that this is "[o]wing to lack of sufficient data". |
| Li et al. [68] | Wuhan City, Hubei-ex-Wuhan, and six provinces, China | Excluded due to new information about the sensitivity of the Wondfo antibody test Silveira et al. [125]. |
| Murhekar et al. (B) [87] | India | Duplicate ([86] report the results from the same study). |
| Pérez-Olmeda et al. [97] | Spain | Duplicate ([102] report the results from the same study). |
| Poustchi et al. [103] | 18 cities of Iran | Death data not found for the target population. |
| Qutob et al. [107] | Palestinian population residing in the West Bank | Not included in Karlinsky and Kobak [57]'s analysis. |
| Ramaswamy et al. [109] | Jabalpur Municipal Corporation ("City of Jabalpur") | While official death numbers are available for the District of Jabalpur (from the "Official Account of the Directorate of Health Services, Government of Madhya Pradesh. (e.g., https://twitter.com/healthminmp/status/1346107670869155843/photo/1; accessed July 27, 2021)), we could not find data for the City of Jabalpur. |
| Silveira et al. [124] | Regions of Brazil | Excluded due to new information about the sensitivity of the Wondfo antibody test [125]. |
| Stefanelli et al. [131] | Five municipalities of the Autonomous Province of Trento, Italy | Convenience sampling. |
| Stringhini et al. (B) [134] | Geneva, Switzerland | Duplicate ([112] report the results from the same study). |
| Truc et al. [138] | Municipality of Cogne, Italy | 95% uncertainty interval not provided for the prevalence estimate. |
| Ulyte et al. [139] | Zurich, Switzerland | Sample not representative of overall population: only school children. |

### 6.3 Details on mortality and covariate data

• For Álvarez-Antonio et al. [3] (“Iquitos, Peru”) (see also Álvarez-Antonio et al. [2]), we used official (post-audit) numbers from the Peru Ministry of Health (MINSA) (see https://www.datosabiertos.gob.pe/dataset/fallecidos-por-covid-19-ministerio-de-salud-minsa/resource/4b7636f3-5f0c-4404-8526) for mortality numbers. We included deaths recorded for 4 districts (”SAN JUAN BAUTISTA”, ”BELEN”, ”PUNCHANA”, and ”IQUITOS”) in the Loreto department. Note that recording the accurate number of deaths is perhaps particularly challenging in Iquitos; see Fraser [36]. In order to acknowledge that there is substantial uncertainty in the post-audit mortality numbers, we widened the interval by lowering the lower bound by 10% and increasing the upper bound by 10%. Limited information is available about mortality amongst LTC residents [142]. As such, no LTC-adjustments were made to the mortality numbers. Data for the pro-portion aged over 65 is from a 2004 World Health Organization report (see https://www.who.int/ageing/projects/intra/phase_two/alc_intra2_cp_peru.pdf; accessed August 2, 2021). GDP data is from the “USD per head, current prices, current PPP” value listed by the Organisation for Economic Co-operation and Development’s Gross Domestic Product, Large regions TL2 database (“OECD database”; see https://stats.oecd.org/, accessed August 10, 2021) for the Loreto region (PE16) . Population data is from the published article [2].
• For Warszawski et al. [146] (“Metropolitan, France”), we obtained mortality data from the French government COVID-19 dashboard (see https://dashboard.covid19.data.gouv.fr/vue-d-ensemble?location=FRA; accessed on August 4, 2021), summing up the cumulative deaths for the 13 regions of Metropolitan France (i.e., European France). (235+394+1616+407+471+6710+3256+959+371+1562+469+836+56 and 257+427+1804+461+528+7358+3518+1032+411+1721+508+927+59). Note that this death total does not include deaths that occurred in LTC facilities. These are listed separately on the French government COVID-19 dashboard under “EHPAD et EMS”. Data for the proportion aged over 65 is for France from 2019 as listed by World Bank’s World Development Indicators (WDI) data (https://data.worldbank.org/indicator/SP.POP.65UP.TO.ZS?locations=FR; accessed August 4, 2021). GDP data is from the OECD database. Population data is from INSEE in report titled “Population totale par sexe et âge au 1er janvier 2020, France métropolitaine” (see https://www.insee.fr/fr/statistiques/1892088?sommaire=1912926; accessed August 4, 2021).
• For Bajema et al. [9] (“DeKalb and Fulton, GA, USA (B)”), we obtained the number of deaths for DeKalb, and Fulton counties from the county-level COVID-19 dataset curated by the New York Times available at: github.com/nytimes/covid-19-data (accessed on April 28, 2021). According to the New York Times analysis, (see: https://web.archive.org/web/20200627101803/https://www.nytimes.com/interactive/2020/us/coronavirus-nursing-homes.html) about 45% of COVID-19 deaths in the state of Georgia were among nursing home residents (as of June 27, 2020). We therefore adjusted the numbers by 45% and obtained 122 (=221-0.45*221) and 136 (=247-0.45*247). Data for the proportion aged over 65 is from https://github.com/JieYingWu/COVID-19_US_County-level_Summaries/blob/master/data/README.md (accessed on April 29, 2021). GDP value is from value listed in the OECD database for the state of Georgia, USA (US13). Population is from the United States census (https://www.census.gov/quickfacts/fultoncountygeorgia and https://www.census.gov/quickfacts/dekalbcountygeorgia ; accessed July 28, 2021) (i.e., 759297 + 1063937). Note that both Bajema et al. [9] and Biggs et al. [18] report the results from the same seroprevalence study.
• For Barchuk et al. [11] (“Saint Petersburg, Russia”), we used excess death numbers as reported by Kobak [61] for the upper bound and official government numbers for the lower bound. Russian official government statistics appear to underestimate the true number of fatalities by a substantial factor [57]. No information was found for mortality amongst LTC residents in Russia; however see: https://www.hrw.org/news/2020/06/02/russia-publish-data-about-covid-19-institutional-care. Data for the pro-portion aged over 65 is from Barchuk et al. [11] (see Table A3 “KOUZh-2018”). GDP value is from OECD database for the “Federal City of Saint Petersburg” (RU29) region. The population value is from https://en.wikipedia.org/wiki/Saint_Petersburg (accessed June 21, 2021)).
• For Biggs et al. [18] (“DeKalb and Fulton, GA, USA (A)”), the number of deaths for DeKalb, and Fulton counties was obtained from the county-level COVID-19 dataset curated by the New York Times available at: github.com/nytimes/covid-19-data (accessed on April 28, 2021). Data for the proportion aged over 65 is from https://github.com/JieYingWu/COVID-19_US_County-level_Summaries/blob/master/data/README.md (accessed on April 29, 2021). According to the New York Times analysis, (see: https://web.archive.org/web/20200627101803/ https://www.nytimes.com/interactive/2020/us/coronavirus-nursing-homes.html; accessed Oct. 12, 2021) about 45% of COVID-19 deaths in the state of Georgia were among nursing home residents (as of June 27, 2020). We adjusted the numbers by 45% and obtained 122 (=221-0.45*221) and 136 (=247-0.45*247). Data for the proportion aged over 65 is from https://github.com/JieYingWu/COVID-19_US_County-level_Summaries/blob/master/data/README.md (accessed on April 29, 2021). GDP value is from value listed in the OECD database for the state of Georgia, USA (US13). Population is from the United States census (https://www.census.gov/quickfacts/fultoncountygeorgia and https://www.census.gov/quickfacts/dekalbcountygeorgia; accessed July 28, 2021) (i.e., 759297 + 1063937). Note that both Bajema et al. [9] and Biggs et al. [18] report the results from the same seroprevalence study.
• For Bruckner et al. [24] (“Orange County, CA, USA”), we obtained the number of cumulative deaths for Orange County from the Orange County Public Works (as referenced by Bruckner et al. [24]) at: data-ocpw.opendata.arcgis.com/datasets/2ec9342ffc814cf58161b1cca57365fd_0 (accessed on April 28, 2021). Orange County Public Works reports deaths of skilled nursing facility (SNF) residents separately. For 2020-07-24, there were a total of 759 deaths including 247 deaths of SNF residents. For 2020-08-30, there were a total of 1209 deaths including 370 deaths of SNF residents. We recorded mortality numbers accordingly. Data for the proportion aged over 65 is from https://github.com/JieYingWu/COVID-19_US_County-level_Summaries/blob/master/data/README.md (accessed on April 29, 2021). GDP value is from value listed for the state of California, USA (US06) from the OECD database. Population value obtained from the United States census (https://www.census.gov/quickfacts/orangecountycalifornia; accessed June 27, 2021).
• For Carrat et al. [27] (“^^^Ile-de-France, France”), we only consider ^^^Ile-de-France phase of the study (see Supplementary Table 1 in Carrat et al. [27] for sampling dates). Data for the number of deaths for ^^^Ile-de-France was obtained from the French government COVID-19 dashboard (see https://dashboard.covid19.data.gouv.fr/vue-d-ensemble?location=FRA; accessed on August 4, 2021). Note that this death total does not include deaths that occurred in long term care facilities (“EHPAD et EMS”). Data for the proportion aged over 65 is from https://contrevues.paris/ile-de-france-comment-le-vieillissement-de-la-population-impacte-lhabitat/ (accessed on April 29, 2021). GDP value is from value listed for ^^^Ile-de-France region (FR1) from OECD database. Population data from the Institut national de la statistique et des etudes economiques (INSEE) (see https://www.insee.fr/fr/statistiques/5002478; accessed July 29, 2021).
• For Chan et al. [28] (“Rhode Island, USA”), we obtained mortality data from the Rhode Island Department of Health available at https://ri-department-of-health-covid-19-data-rihealth.hub.arcgis.com/ (accessed July 28, 2021). Note that Chan et al. [28] state that: “As of May 31, Rhode Island had reported 827 cumulative lab-confirmed SARS-CoV-2–involved deaths; 78.5% of these deaths were associated with congregate care facilities. We therefore reduced the mortality numbers by 78.5% (= 684×(1−0.785) and = 877×(1−0.785)). Data for the proportion aged over 65 is from the U.S. Department of Commerce https://www.census.gov/quickfacts/RI (accessed on June 21, 2021). GDP value is from value listed for the state of Rhode Island, USA (US44) in the OECD database. The population value is obtained from the United States census (https://www.census.gov/quickfacts/RI; accessed July 28, 2021).
• For Govern d’Andorra [43] (“Andorra”) (see also Royo-Cebrecos et al. [115]), we obtained mortality numbers from the Government of Andorra (https://www.govern.ad/coronavirus; accessed on June 28, 2021) as tabulated by Wikipedia (https://en.wikipedia.org/wiki/COVID-19_pandemic_in_Andorra; accessed on June 28, 2021). We substracted 31.45% of deaths (35 = 51-0.3145*51 for both lower and upper bounds) following the reporting of Carrasco et al. [26] which found that 39 out of 124 COVID-19 deaths in Andorra (for the period between March, 2020 and April, 2021) were amongst residents in nursing homes. Carrasco et al. [26] note that: “218/354 (61.6%) residents in nursing homes were infected and the fatality rate was 39/218 (17.8%).” Data for the proportion aged over 65 is from https://www.indexmundi.com/andorra/demographics_profile.html (accessed on June 28, 2021) and GDP data is from The World Factbook (https://statisticstimes.com/economy/countries-by-gdp-capita-ppp.php; accessed on June 28, 2021)). The population number is as stated in the Royo-Cebrecos et al. [115] which is the published article reporting on the study.
• For Kar et al. [56] (“Puducherry District, India”), data for the number of deaths for the Puducherry District was obtained from https://covid19dashboard.py.gov.in/Reporting/District (accessed on August 5, 2021). We multiplied the number recorded for 14 days after the end of the sampling window by a factor of 2.1 (based on the upper bound of Mukherjee et al. [83]’s estimated underreporting factor for Puducherry) in order to account for potential underreporting. As such, our interval is relatively wide and reflects the substantial uncertainty in the true number of deaths. There is insufficient information about deaths of LTC residents in India to make any LTC adjustments; see Jayeeta Rajagopalan and Alladi [55]. Data for the proportion aged over 65 is from http://statisticstimes.com/demographics/india/puducherry-population.php (accessed on June 22, 2021). GDP value is from value listed for Puducherry (IN34) from OECD database. Population for Puducherry District from Kar et al. [56] who note that: “Puducherry district, population *≈*1.25 million, is located in southern India.”
• For Khalagi et al. [58] (“Iran”), Karlinsky and Kobak [57] suggest that official mortality data for Iran is highly inaccurate and suggest that there have been approx. 58,092 excess deaths in Iran for the period until Sept. 21, 2020. This coincides with the estimate provided by ghafari2020excess of 58,900 (95% CI 46,900 - 69,500) for the period from December 22, 2019 to September 21, 2020. Excess mortality data is only available in quarterly format from Kobak [61]. Therefore it is not possible to obtain mortality data for the specific dates we require: 2020-08-17 and 2020-11-14. Official statistics for these dates are 19,804 and 41,034 (see https://en.wikipedia.org/wiki/COVID-19_pandemic_in_Iran; accessed on June 28, 2021, which cites the Iranian Ministry of Health and Medical Education). We used 19,804 for the lower bound and multiplied 41,034 by Kobak [61]’s estimated undercount ratio of 2.4 to obtain an upper bound of 98071. (see https://www.ncbi.nlm.nih.gov/pmc/articles/PMC7852240/pdf/nihpp-2021.01.27.21250604v3.pdf; accessed on June 28, 2021). In Iran, LTC is provided mostly informally by family caregivers [41]. As such, no adjustments were made to exclude deaths of LTC residents. Data for the population and the proportion of the population aged over 65 is from the World Bank World Development Indicators (WDI) data (https://data.worldbank.org/indicator/SP.POP.65UP.TO.ZS?locations=IR; accessed on June 28, 2021). GDP value is also from the WDI data (see: https://data.worldbank.org/indicator/NY.GDP.PCAP.PP.CD (last updated 2021-04-26)).
• For Mahajan et al. [74] (“Connecticut, USA”), mortality data obtained from Wikipedia (https://en.wikipedia.org/wiki/COVID-19_pandemic_in_Connecticut; accessed on July 22, 2021), who reference (https://portal.ct.gov/Coronavirus). Mortality numbers (4,287 and 4,450) are multiplied by 26.5% (i.e. by 1079/4071) due to the fact that approx. 26.5% of COVID-19 deaths in Connecticut occurred in the community-dwelling population (i.e., “non-congregate” population) according to Mahajan et al. [73]. Data for the proportion aged over 65 is derived from Table 1 of Mahajan et al. [73]. GDP value is from value listed for the state of Connecticut, USA (US09) from the OECD database. Population value is obtained from [73].
• For Malani et al. [76] (“Tamil Nadu, India”), the population of Tamil-Nadu, India, was obtained from https://www.indiacensus.net/states/tamil-nadu (accessed July 27, 2021). Mortality data for the number of deaths for Tamil Nadu was obtained from the Tamil Nadu Government (see https://stopcorona.tn.gov.in/archive/; accessed June 24, 2021). We multiplied the number recorded for 14 days after the end of the sampling window of 11,909 by a factor of 2.5 (based on the upper bound of Mukherjee et al. [83]’s estimated underreporting factor for Tamil Nadu). There is insufficient information about deaths of LTC residents in India to make any LTC adjustments; see Jayeeta Rajagopalan and Alladi [55]. GDP value is from value listed for Tamil-Nadu (IN33) from the OECD database. Data for the proportion aged over 65 is from https://statisticstimes.com/demographics/ india/tamil-nadu-population.php (accessed July 27, 2021).
• For Melotti et al. [79] (“Gardena Valley, Italy”), we obtained information from the report “Study on the prevalence of Covid-19 in the Val Gardena - June 2020” (see: https://astat.provincia.bz.it/it/news-pubblicazioni-info.asp?news_action=300&news_image_id=1074603; accessed July 27, 2021). In this report, it is noted that ”the number of deaths from this disease in 17 people…”. Stefano Lombardo (in a personal communication) notes that the source of this number is Astat, Bolzano (”statistics on deaths by cause”) and has issued a correction (via personal communication) to indicate that the number should be 20 (”In spring 2020 the covid19-deaths in Gardena were estimated at 17; at the end of the year the official statistics closed at 20 (march+april).”) In a further personal communication (”R: R: INFERRING THE COVID-19 IFR”; Oct. 18, 2021), Lombardo clarified that out of these 20 deaths 3 occurred at home, 12 at the hospital, and 5 at a nursing home. The study looks specifically at three municipalities (Ortisei, Santa Cristina, and Selva) and we used age data from https://www.citypopulation.de/en/italy/trentinoaltoadige/bolzano/021089selva_di_val_gardena/ and http://italia.indettaglio.it/eng/trentinoaltoadige/ortisei.html (accessed August 4, 2021). GDP value is from OECD database for the Province of Bolzano-Bozen (ITH1).
• For Ministry of Health of Israel [81] (“Israel”) (see also Reicher et al. [110]), mortality data obtained from https://en.wikipedia.org/wiki/COVID-19_pandemic_in_Israel (accessed July 27, 2021) which references Israel’s Ministry of Health’ official coronavirus updates, it’s Telegram channel, and Israel’s Corona National Information and Knowledge Center. According to Sharona Tsadok-Rosenbluth [121], a large porpotion of COVID-19 deaths in Israel occurred in long-term care facilities (LTCFs). Specifically, by the end of July, 2020 (2020/07/28), the percentage of deaths that occurred in LTCFs was 51%, and by the end of September, 2020 (2020/09/29), the percentage of deaths that occurred in LTCFs was 40%. Based on these figures we adjusted the death numbers 382 and 1659 to: 187 (=382-0.51*382) and 995 (=1659-0.4*1659). Data for the proportion aged over 65 is from https://brookdale.jdc.org.il/wp-content/uploads/2018/02/MJB-Facts_and_Figures_Elderly-65_in_Israel-2018_English.pdf (accessed July 27, 2021) and GDP value is from OECD. Population for Israel is from World Bank’s World Development Indicators (WDI) data. Note that while [6] categorizes the sampling method of the Ministry of Health of Israel [81] study as “stratified probability,” Reicher et al. [110] note that the study used blood samples from “insured individuals who arrived at the HMOs [Health Maintenance Organizations] to undergo a blood test for any reason” and caution that, as a consequence, the “study might not reliably represent the entire population.”
• For Murhekar et al. [86] (“India”), we obtained the number of cumulative deaths for India, from the Our World in Data COVID-19 dataset available at: ourworldindata.org/coronavirus/country/india (accessed on April 28, 2021). We multiplied the number recorded for 14 days after the end of the sampling window of 12,573 by a factor of 3.64 (based on the upper bound of Purkayastha et al. [105]’s estimated underreporting factor for India (first wave)) in order to account for potential underreporting. As such, our interval is relatively wide and reflects the uncertainty in the true number of deaths: [4172, 45766]. (Purkayastha et al. [105]: “Estimates from epidemiological models: For wave 1 our estimate… [is] an underreporting factor for cases estimated at 11.11 (95% CrI 10.71 – 11.47) and for deaths at 3.56 (95% CrI 3.48 – 3.64).”) There is insufficient information about deaths of LTC residents in India to make any LTC adjustments; see Jayeeta Rajagopalan and Alladi [55]. Data for the proportion aged over 65 is from World Bank’s World Development Indicators (WDI) data (see https://data.worldbank.org/indicator/SP.POP.65UP. TO.ZS (last updated 2021-03-19)). GDP value is from OECD database. Population value is from the World Bank (see https://data.worldbank.org/indicator/SP.POP.TOTL?locations=IN; accessed on April 28, 2021).
• For Nawa et al. [89], we concluded that 0 deaths had occurred based on official records for the Tochigi prefecture (see https://www.nippon.com/en/japan-data/h00657/; accessed July 12th, 2021). For the infection interval we use the lowest bound of the confidence interval given for the “unimputed dataset” and the upper bound of the confidence interval given for the “imputed dataset” from Table 1 of Nawa et al. [89]. Note that among the 181 participants in the Nawa et al. [89] study who were aged 65 years or older none were positive (0/181). This suggests that the prevalence amongst those at greatest risk of death (i.e., amongst the elderly) may be lower than in the greater population. However, since the Nawa et al. [89] found only 3 positive cases out of 742 individuals tested, inference on this is limited. Also note that, as of July 13, 2020, amongst the 36 individuals who tested positive by pcr-testing in Utsunomiya City, 4 were in their 70s and none were older than 80 (see https://web.archive.org/web/20200714014725/https://www.city.utsunomiya.tochigi.jp/kurashi/kenko/kansensho/etc/1023506.html). Data for proportion aged over 65 is from https://www8.cao.go.jp/kourei/english/annualreport/2017/pdf/c1-1.pdf (accessed July 12th, 2021). GDP value is from OECD database for Northern-Kanto, Koshin (JPC) region. Population value is from https://www.city.utsunomiya.tochigi.jp/shisei/gaiyo/1007461.html (accessed July 12th).
• For Office of National Statistics [92] (“England, UK (A)”), we used numbers obtained from the Office of National Statistics, dataset (“deaths registered weekly in England and Wales”; see publishedweek532020.xlsx, sheet “UK - Covid-19 - Weekly occurrences”) retrieved from https://www.ons.gov.uk/peoplepopulationandcommunity/birthsdeathsandmarriages/deaths/datasets/weeklyprovisionalfiguresondeathsregisteredinenglandandwales (accessed Oct. 12, 2021). We subtracted those deaths attributed to Wales, and subtracted deaths attributed to nursing home residents as recorded in the “Deaths involving COVID-19 in the care sector, England and Wales” dataset (available at https://www.ons.gov.uk/peoplepopulationandcommunity/birthsdeathsandmarriages/deaths/datasets/deathsinvolvingcovid19inthecaresectorenglandandwales; (accessed Oct. 12, 2021)). For 2020-05-15 (the closest available date to 2020-07-14), there were 41135 total deaths for England (= 43168 (England and Wales) - 2033 (Wales)) from which we subtracted 15160 deaths of care home residents (“England, ONS Data”) to obtain 25975. For 2020-09-25 (the closest available date to 2020-09-26), there were 50589 total deaths for England from which we subtracted 19880 deaths of care home residents (“England, ONS Data”) to obtain 30709. Data for the proportion aged over 65 is for England from 2019 as listed by LG Inform (see https://tinyurl.com/4vhrb2uu; accessed on April 29, 2021). GDP value is obtained by taking the average of values listed in the OECD database for the regions of “South West England”, “North East England”, “East of England”, “North West England”, and “South East England”. Population value is from the Office of National Statistics report: “Population estimates for the UK, England and Wales, Scotland and Northern Ireland: mid-2020” (see Table 2) (https://www.ons.gov.uk/peoplepopulationandcommunity/populationandmigration/populationestimates/bulletins/annualmidyearpopulationestimates/mid2020; accessed July 30, 2021).
• For Pagani et al. [93] (“Castiglione d’Adda, Italy”), data was taken directly from the paper. Pagani et al. [93] does not cite a specific source for deaths but notes: “From the 1st of January to the 31st of March 2020, 76 deaths (1.65% of the population) have been recorded in CdA, of which 47 were officially attributed to COVID-19.” We have not been able to verify these numbers using a publicly available dataset. With regards top LTC deaths, Costantino Pesatori, the mayor of Castiglione D’Adda, is quoted as saying: “In our retirement home alone, we have had at least seven casualties.” (“Sono morti tanti anziani, è vero. Solo nella nostra casa di riposo abbiamo avuto almeno sette perdite.” See La Stampa: https://www.lastampa.it/topnews/primo-piano/2020/03/16/news/la-strage-silenziosa-di-castiglione-d-adda-43-morti-in-tre-settimane-1.38599650; accessed Oct. 12, 2021). With this in mind, we subtracted 7 and 8 deaths from the total of 47 cited in the paper. Note that it appears that 100% of residents in the nursing home were evntually infected (see https://www.ilcittadino.it/stories/Cronaca/covid-casa-di-riposo-di-castiglione-tutti-gli-anziani-contagiati_61445_96/). For population, Pagani et al. [93] states that “Castiglione d’Adda (CdA) is a town of 4605 inhabitants (according to data from the local registry office at the time of our study) […]”. Data for the proportion aged over 65 is from https://www.citypopulation.de/en/italy/localities/lombardia/lodi/09801410001castiglione_dadda/ (accessed July 27, 2021). GDP value is from OECD database for Lombardy region (ITC4).
• For Petersen et al. [98] (“Faroe Islands, Denmark”), information on deaths for the Faroe Islands was obtained from corona.fo/hagtol, the government information website concerning COVID19 in the Faroe Islands, which indicated zero deaths. GDP value is from value listed for Denmark from OECD database. Data for the proportion aged over 65 is from Index mundi (see https://tinyurl.com/bb2pwrx6; accessed on April 29, 2021) which cites the CIA World Factbook as a source. Population value is from Petersen et al. [98] (“In the Faroe Islands, a geographic isolate of 52,154 inhabitants.”).
• For Pollán et al. [102] (“Spain”), data for the number of deaths was obtained from Wikipedia (en.wikipedia.org/wiki/COVID-19_pandemic_in_Spain; accessed on April 28, 2021) which sourced the information from the Centro Nacional de Epidemiologa (cnecovid.isciii.es/covid19/). Note that, the number of deaths of for 2020-05-11 of 26,920 (14 days after the start of the sampling window), is actually higher than the number of deaths for 2020-05-25 of 26,834 (14 days after the end of the sampling window). This may be due to a reporting issue which is noted by Wikipedia: “Figures for 2020-05-24 to 2020-06-17 include corrections in the validation of past data from several autonomous communities as a result of the transition to a new surveillance methodology implemented from 2020-05-11.” We subtracted 34% from the mortality numbers (26834, and 26920) to account for those deaths that occurred in long-term care facilities. This follows the practice done by Pastor-Barriuso et al. [94] who subtracted 9,909 deaths from a the total of 29,137; see discussion within Pastor-Barriuso et al. [94] for details. GDP value is from value listed for Spain from OECD database. Data for the proportion aged over 65 is from World Bank’s World Development Indicators (WDI) data (see https://data.worldbank.org/indicator/SP.POP.65UP.TO.ZS (last updated 2021-03-19)). Population value is from the World Bank for 2020 (https://data.worldbank.org/indicator/SP.POP.TOTL?locations=ES; accessed July 30, 2021).
• For Radon et al. [108] (“Munich, Germany”), the number of deaths was obtained from the official database of the city of Munich (see https://www.muenchen.de/rathaus/Stadtinfos/Coronavirus-Fallzahlen.html; accessed June 22, 2021). Gleich et al. [42] conclude that: “One quarter of all COVID-19 deaths in Munich occurred in the context of nosocomial outbreaks in elderly, chronically ill residents of nursing facilities.” Based on this finding, we subtract 25% from the lower bound (292) and the upper bound (1040) to obtain the numbers: 219 and 780. Population data is from https://web.archive.org/web/20201218195500/https://www.muenchen.de/rathaus/Stadtinfos/Statistik/Bev-lkerung.html (accessed June 22, 2021) and data for those aged over 65 from https://www.muenchen.de/rathaus/dam/jcr:459cce24-3894-4d4d-b144-4185b750a310/LHM.Stat_Faltkarte_2019_englisch.pdf (accessed June 22, 2021). Finally, GDP value is from value listed for Bavaria (GE2) from the OECD database.
• For Reyes-Vega et al. [111] (“Lima, Peru”), note that Peru audited death records and their data was updated on May 31, 2021; see Dyer [32] for details. We used official (post-audit) numbers from the Peru Ministry of Health (MINSA) (see https://www.datosabiertos.gob.pe/dataset/fallecidos-por-covid-19-ministerio-de-salud-minsa/resource/4b7636f3-5f0c-4404-8526; accessed June 22, 2021) for mortality numbers. Note that the target population is Lima Metropolitana, an area that includes both area that includes Peruvian provinces of Lima and Callao. As such, we sum deaths recorded for both Lima and Callao provinces. In order to acknowledge that there is substantial uncertainty in the post-audit mortality numbers, we widened the interval by lowering the lower bound by 10% and increasing the upper bound by 10%. Limited information is available about mortality amongst LTC residents [142]. As such, no LTC-adjustments were made to the mortality numbers. GDP value is from value listed for Lima (PE15) from the OECD database. Data for the population aged over 65 from https://www.citypopulation.de/en/peru/admin/15lima/ (accessed July 27, 2021). Population value obtained from https://www.minsa.gob.pe/reunis/data/poblacion_estimada.asp (accessed July 27, 2021). Note that Reyes-Vega et al. [111] state that: “The study area has an estimated 10.7 million inhabitants.”
• For Richard et al. [112] (“Geneva, Switzerland (B)”), note that this study is excluded from the Chen et al.-based analysis set but included in the Serotracker-based analysis set. For Richard et al. [112], mortality data for the canton of Geneva were obtained from an excel file made publicly available by a Swiss government website at: ge.ch/document/covid-19-donnees-completes-debut-pandemie (accessed on April 28, 2021). According to reporting from mid-Jun 2020 by the canton of Geneva (Département de la sécurité, de l’emploi et de la santé (DSES) - Direction générale de la santé - Service du médecin cantonal), about 45% of deaths were among nursing home residents (“résidents des établissements médico-sociaux (EMS)”); see document at https://www.ge.ch/document/19696/annexe/61. We therefore adjusted the numbers by 45%: 122 (=222-0.45*222) and 156 (= 283-0.45*283). Data for the population and the proportion aged over 65 is from https://www.bfs.admin.ch/bfs/en/home/statistics/regional-statistics/regional-portraits-key-figures/cantons/geneva.html (accessed on April 28, 2021). GDP value is from value listed for Lake Geneva Region (CH01) from the OECD database.
• For Samore et al. [116] (“Four counties in UT, USA”), data for the number of deaths for the counties of Utah county, Salt Lake county, Davis county, and Summit county, was obtained from the county-level COVID-19 dataset curated by the New York Times available at: github.com/nytimes/covid-19-data (accessed on April 28, 2021). The Salt Lake Tribune reported on July 14, 2020 that 42% of COVID-19 deaths in Utah had been attributed to nursing homes (“According to the state health department, 95 of Utah’s 226 COVID-19 deaths have been attributed to the facilities.”; see https://www.sltrib.com/news/2020/07/14/utah-nursing-homes-keep/; accessed Oct. 12, 2021). This agrees with analysis by the New York Times (see: https://web.archive.org/web/20200627101803/ https://www.nytimes.com/interactive/2020/us/coronavirus-nursing-homes.html; accessed Oct. 12, 2021). We therefore adjusted the mortality number by 42% and obtained 40 (=69-69*0.42) and 97 (=168-168*0.42). Data for the proportion aged over 65 is from https://github.com/JieYingWu/COVID-19_US_County-level_Summaries/blob/master/data/README.md (accessed on April 29, 2021). GDP value is from value listed for the state of Utah, USA (US49) in the OECD database. Population value is from US county-level census data for 2019 for the four counties (636235+1160437+355481+42145) (see https://www.census.gov/data/datasets/time-series/demo/popest/2010s-counties-total.html; accessed July 30, 2021).
• For Santos-Hö vener et al. [117] (“Kupferzell, Germany”), data for the number of deaths for Kupferzell, Germany was obtained directly from Santos-Hö vener et al. [117] which cites the Robert Koch Institute. Despite efforts, no publicly available dataset was found which could confirm these numbers specific these numbers. Santos-Hö vener et al. [117] report that 3 deaths occurred in the town for the period before July, 2020, and do not specify if these were nursing home residents. Santos-Hö vener et al. [117] do however specify the ages of those who died: “aged 59, 81 and 91 years.” Data for the proportion aged over 65 is from: https://ugeo.urbistat.com/AdminStat/en/de/demografia/eta/kupferzell/20172564/4 (accessed on April 29, 2021). GDP value is from value listed for Baden-Württemberg (DE1) from the OECD database. Population value is from Wikipedia (https://als.wikipedia.org/wiki/Kupferzell; accessed July 30, 2021) which cites the Statistisches Landesamt Baden-Wü rttemberg – Bevö lkerung nach Nationalität und Geschlecht am 31. Dezember 2019 (CSV-Datei).
• For Selvaraju et al. [118] (“Chennai, Tamil Nadu, India”), mortality data for the number of deaths for Chennai were obtained from the Greater Chennai Corporation (see http://covid19.chennaicorporation.gov.in/; specifically, numbers published for July 15, 2020 and August 14, 2020 were listed at: https://twitter.com/chennaicorp/status/1283280173132128259 and https://twitter.com/chennaicorp/status/1294140456868052993; accessed June 22, 2021). We multiplied the number recorded for 14 days after the end of the sampling window of 2,384 by a factor of 2.5 (based the upper bound of Mukherjee et al. [83]’s estimated underreporting factor for Tamil Nadu) in order to account for potential underreporting. Note that Ariel Karlinsky has assembled excess mortality data for Chennai (see: https://github.com/akarlinsky/world_mortality/tree/main/local_mortality; accessed August 4, 2021). These data, taking into account the weekly numbers and five-year pre-pandemic trend, suggest an interval of [1344, 2454]. However, we prefer not to use this much narrower interval since the excess mortality data have not been adjusted to account for the potentially substantial underreporting of all-cause deaths (see discussion in Anand et al. [4]). There is insufficient information about deaths of LTC residents in India to make any LTC adjustments; see Jayeeta Rajagopalan and Alladi [55]. Data for the proportion aged over 65 is from: https://statisticstimes.com/demographics/india/tamil-nadu-population.php (accessed June 22, 2021). GDP value is from value listed for Tamil Nadu (IN33) from the OECD database. Population value is from https://www.populationu.com/cities/chennai-population (accessed June 22, 2021).
• For Sharma et al. [120] (“Delhi, India”), infection rate estimates are based on survey data from round 1 of the study (August 1-7). Data for the number of deaths for Delhi was obtained from Wikipedia (en.wikipedia.org/wiki/COVID-19_pandemic_in_Delhi; accessed on April 28, 2021) which sourced the information from the Delhi State Health Bulletin (https://delhifightscorona.in/). We multiplied the number recorded for 14 days after the end of the sampling window of 4,270 by a factor of 6.3 (based on Mukherjee et al. [83]’s estimated underreporting factor) in order to account for potential underreporting. As such, our interval is relatively wide and should reflect the uncertainty in the true number of deaths: [4,188, 26,901]. There is insufficient information about deaths of LTC residents in India to make any LTC adjustments; see Jayeeta Rajagopalan and Alladi [55]. Data for the proportion aged over 65 is from Statistics Times for 2011 (http://statisticstimes.com/demographics/india/delhi-population.php; accessed on April 29, 2021). GDP value is from value listed for the National Capital Territory of Delhi (IN03) from the OECD database. Note that we use 18.9 million as the estimated population of Delhi, the number cited by both Sharma et al. [120] and Bhattacharyya et al. [17], while Chen et al. [29] use a much different number: 30,290,936 (see Table S14 in Supplementary Appendix 2 of Chen et al. [29]).
• Snoeck et al. [126] (“Luxembourg”) “recruited a representative sample of the Luxembourgish population” between April 16th and May 5th, and obtained a 95% CI of [1.23%, 2.77%]. Two different 95% CIs, obtained with and without adjustment for age, gender and canton are provided in the paper: [1.23%; 2.67%] and [1.34%; 2.77%]. As such, we record [1.23%; 2.77%] for our IR interval. Data for the number of deaths was obtained from the Our World in Data COVID-19 dataset available at: ourworldindata.org/coronavirus/country/luxembourg (accessed on April 28, 2021). Hansen and Dalesio [46] report that: “Of the 507 fatalities from Covid-19 in 2020, 241 had lived in housing offering care for the elderly, the Health Ministry reported to Parliament in January.” As such we adjusted the numbers by subtracting 47% to obtain 47 (=89-0.47*89) and 58 (=109-0.47*109). GDP value is from value listed for Luxembourg (LU00) from the OECD database. Data for the proportion aged over 65 is from World Bank’s World Development Indicators (WDI) data (see https://data.worldbank.org/indicator/SP.POP.65UP.TO.ZS (last updated 2021-03-19)). Population value from World Bank for Luxembourg, 2020 (see https://data.worldbank.org/indicator/SP.POP.TOTL?locations=LU; accessed July 30, 2021). We note that, while Arora et al. [6] categorizes the sampling method for the Snoeck et al. [126] study as “stratified probability,” Snoeck et al. [126] explains that “the sample of participants was enrolled through the use of a non-probabilistic web panel” and caution that “[t]here are pros and cons of using web panels for surveys,” specifically that “online surveys may be biased samples because the respondents are self-selected.” Levin et al. [66] specifically identify Snoeck et al. [126] as an example of a “active recruitment” study.
• For Sood et al. [127] (“Los Angeles County, CA, USA”), data for the number of deaths for Los Angeles County, CA was obtained from the County of Los Angeles Public Health for 2020-04-24 (913 total deaths, and 384 deaths in institutional settings “as of 8pm 4/26”; see https://web.archive.org/web/20200426230946/http://publichealth.lacounty.gov/media/coronavirus/locations.htm; accessed Oct. 12, 2021) and for 2020-04-28 (1111 total deaths, and 505 in institutional settings “As of 8:00pm 4/29”; see https://web.archive.org/web/20200501065125/ http://publichealth.lacounty.gov/media/coronavirus/locations.htm; accessed Oct. 12, 2021). To be clear, the County of Los Angeles Public Health reports deaths in institutional settings separately. Sood et al. [127] notes that “Residents of Los Angeles County, California, within a 15-mile (24 km) radius of the testing site were eligible for participation.” Data for the proportion aged over 65 is from https://github.com/JieYingWu/COVID-19_US_County-level_Summaries/blob/master/data/README.md (accessed on April 29, 2021). GDP value is from value listed for the state of California, USA, (US06) in the OECD database. Population value is from US county-level census data for 2019 (see https://www.census.gov/data/datasets/time-series/demo/popest/2010s-counties-total.html; accessed July 30, 2021).
• For Statistics Jersey [129] (“Jersey, UK (A)”), data for the number of deaths was obtained from the Government of Jersey website (https://www.gov.je/datasets/listopendata?listname=COVID19DeathsClassification; accessed on April 28, 2021) summing both “probable COVID-19” deaths and “laboratory proven” COVID-19 deaths. According to reporting by the BBC on Apr. 27 2020 (see https://t.co/2mj8YJbMsL?amp=1; accessed Oct. 12, 2021), 7 out of 19 deaths occurred in care homes. With this in mind, we adjust our mortality numbers (27, and 28) by 37% to obtain 17 and 18. Data for the proportion aged over 65 is from https://www.indexmundi.com/jersey/demographics_profile.html (accessed on April 29, 2021). GDP value is from value listed for United Kingdom (GBR) in the OECD database. Population value is from Statistics Jersey [129].
• For Statistics Jersey [130] (“Jersey, UK (B)”) data for the number of deaths was obtained from the Government of Jersey website (https://www.gov.je/datasets/listopendata?listname=COVID19DeathsClassification; accessed on April 28, 2021) summing both “probable COVID-19” deaths and “laboratory proven” COVID-19 deaths. According to reporting by the BBC on Apr. 27 2020 (see https://t.co/2mj8YJbMsL?amp=1; accessed Oct. 12, 2021), 7 out of 19 deaths occurred in care homes. With this in mind, we adjust our mortality numbers (31, and 31) by 37% to obtain 20 and 20. Data for the proportion aged over 65 is from https://www.indexmundi.com/jersey/demographics_profile.html (accessed on April 29, 2021). GDP value is from value listed for United Kingdom (GBR) in the OECD database. Population value is from Statistics Jersey [130].
• For Streeck et al. [132], the number of deaths for Gangelt, Kreis Heinsberg, Germany, is directly noted in the Streeck et al. [132] article (n=8 deaths). No publicly available dataset was found, however we did find information on the Gangelt municipal bulletin (see www.gangelt.de/news/226-erster-corona-fall-in-nrw; accessed on April 28, 2021) which suggests that 8 is a reasonable number to consider. According to reporting, Gangelt did not suffer substantial (if any) fatalities in LTC facilities during the study period (MedWatch reports that during the study period: “there were no deaths of patients with Covid-19 in the Katharina-Kasper-Heim [nursing home] - nor in a nursing home with 80 residents, which is located in the Breberen district.” see: https://medwatch.de/2020/05/08/wie-manche-virologen-vertrauen-verspielen/; accessed on Oct. 18, 2021). Therefore, we did not subtract any deaths to account for nursing home residents. Note that the seroprevalence estimates for this study are based on the results of antibody tests as well as the results of PCR tests. As such the 14 day offset may not be as appropriate for this study as compared to other studies that consider only antibody tests. Data for the proportion aged over 65 is from Figure S1 of Streeck et al. [132]. GDP value is from value listed for the North Rhine-Westphalia region (DEA) from the OECD database. Population value is from Streeck et al. [132] (“In the German community of Gangelt (12,597 inhabitants, January 1, 2020)”).
• For Stringhini et al. [133] (“Geneva, Switzerland (A)”), mortality data for the canton of Geneva were obtained from an excel file made publicly available by a Swiss government website at: ge.ch/document/covid-19-donnees-completes-debut-pandemie (accessed on April 28, 2021). According to reporting from mid-June 2020 by the canton of Geneva (Département de la sécurité, de l’emploi et de la santé (DSES) - Direction générale de la santé - Service du médecin cantonal), about 45% of deaths were among nursing home residents (“résidents des établissements médico-sociaux (EMS)”); see document at https://www.ge.ch/document/19696/annexe/61. We adjusted the numbers by 45%: 122 (=222-0.45*222) and 155 (= 281-0.45*281). Data for the proportion aged over 65 is from https://www.bfs.admin.ch/bfs/en/home/statistics/regional-statistics/regional-portraits-key-figures/cantons/geneva.html. GDP value is from value listed for Lake Geneva Region (CH01) from the OECD database. Population value is from the Swiss Federal Statistical Office (https://www.bfs.admin.ch/bfs/en/home/statistics/regional-statistics/regional-portraits-key-figures/cantons/geneva.html; accessed July 30, 2021).
• For Vos et al. [141], mortality data for the Netherlands was obtained from the National Institute for Public Health and the Environment https://www.rivm.nl/coronavirus-covid-19/grafieken; accessed Oct. 12, 2021. The numbers used are those listed under “Aantal overledenen naar datum van overlijden” (“Number of deceased by date of death”). Based on available information (see https://www.verenso.nl/nieuws/archief/2020/update-registratie-verpleeghuizen-12-mei-2020 (accessed Oct. 12, 2021)) there were 1696 nursing home residents who died as of May 12, 2020. This represents about 30% of all reported COVID-19 deaths in the country (1696/5738) up until May 12, 2020. As such we will adjust our numbers by 30% : 2621 (=3744-3744*0.30) and 4186 (=5980-5980*0.30). The GDP value is from data listed for Netherlands (NLD) from the OECD database. Data for the proportion aged over 65 is also from World Bank’s World Development Indicators (WDI) data (see https://data.worldbank.org/indicator/SP.POP.65UP.TO.ZS (last updated 2021-03-19)). Population value is from the World Bank (https://data.worldbank.org/indicator/SP.POP.TOTL?locations=NL; accessed July 30, 2021).
• For Ward et al. [145] (“England, UK (B)”), infection rate estimates are based on survey data from from the first survey (20 June - 13 July). We used numbers obtained from the Office of National Statistics, dataset (“deaths registered weekly in England and Wales”; see publishedweek532020.xlsx, sheet “UK - Covid-19 - Weekly occurrences”) retrieved from https://www.ons.gov.uk/peoplepopulationandcommunity/birthsdeathsandmarriages/deaths/datasets/weeklyprovisionalfiguresondeathsregisteredinenglandandwales (accessed Oct. 12, 2021). We subtracted those deaths attributed to Wales, and subtracted deaths attributed to nursing home residents as recorded in the “Deaths involving COVID-19 in the care sector, England and Wales” dataset (available at https://www.ons.gov.uk/peoplepopulationandcommunity/birthsdeathsandmarriages/deaths/datasets/deathsinvolvingcovid19inthecaresectorenglandandwales; (accessed Oct. 12, 2021)). For 2020-07-03 (the closest available date to 2020-07-04), there were 48718 total deaths for England (= 51207 (England and Wales) - 2489 (Wales)) from which we subtracted 19192 deaths of care home residents (“England, ONS Data”) to obtain 29526. For 2020-07-31 (the closest available date to 2020-07-27), there were 49593 total deaths for England from which we subtracted 19555 deaths of care home residents (“England, ONS Data”) to obtain 30038. Note that the numbers of cumulative deaths for England, from the UK coronavirus dashboard (https://coronavirus.data.gov.uk/details/deaths?areaType=nation&areaName=England; accessed on April 29, 2021) are somewhat different. For example, for 2020-07-04, the UK coronavirus dashboard reports 47,608 deaths for England “with COVID-19 in the death certificate” and 36,233 deaths for England that occurred “within 28 days of a positive test.” Data for the proportion aged over 65 is for England from 2019 as listed by LG Inform (see https://tinyurl.com/4vhrb2uu; accessed on April 29, 2021). GDP value is obtained by taking the average of values listed in the OECD database for the regions of ”South West England”, ”North East England” ”East of England”, ”North West England” and ”South East England”. Population value is from the Office of National Statistics report: “Population estimates for the UK, England and Wales, Scotland and Northern Ireland: mid-2020”’ (see Table 2 at: https://www.ons.gov.uk/peoplepopulationandcommunity/populationandmigration/populationestimates/bulletins/annualmidyearpopulationestimates/mid2020; accessed July 30, 2021).
• For Yoshiyama et al. [150] (“Tokyo, Japan”), for the number of deaths for “2020-06-15” and “2020-06-21”, we used official numbers from the Japanese Ministry of Health, Labour, and Welfare (see: https://www.mhlw.go.jp/content/10906000/000640012.pdf and https://www.mhlw.go.jp/content/10906000/000641965.pdf; accessed August 6, 2021). Estévez-Abe and Ide [33] report: “As of May 8, only 14% of COVID-19-related deaths in Japan had occurred in LTCFs (Kyodo News Service, 2020).” We adjusted the numbers accordingly: 270 (=314-0.14*314) and 275 (=320-0.14*320). Data for the population and the proportion of the population aged over 65 is from the Tokyo metropolitan government (https://www.metro.tokyo.lg.jp/tosei/hodohappyo/press/2021/01/28/01.html and https://www.metro.tokyo.lg.jp/ENGLISH/ABOUT/HISTORY/history03.htm; accessed August 6, 2021) and GDP value is obtained by taking the average of values listed in the OECD database for the regions of “Northern-Kanto, Koshin” (JPC) and “Southern-Kanto” (JPD).

### 6.4 MCMC details and R code

Note that, in order to improve the MCMC mixing, we replace the binomial distribution for *CC_k_* as described in (2), with

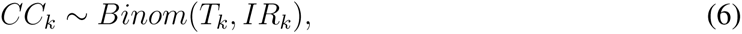

for *k* = 1*, . . . , K*. For any sufficiently large *P_k_* , this simplification will make little to no difference. Then, since the distributions of *C_k_* and *D_k_|C_k_* are both binomials (see (2) and (3)), we have that unconditionally:

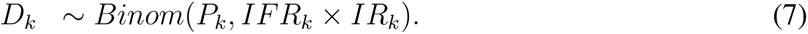

Note that *Z*_1_*_k_* is set equal to the centred and scaled logarithm of 65*yo_k_* , and *Z*_2_*_k_* is set equal to the centred and scaled logarithm of *GDP_k_*.

The following JAGS-code is used in R for the analysis. See the JAGS user manual [101] for details about writing JAGS code (and also Qi et al. [106] for details about the specification of censored data in JAGS and the alternative modeling strategy necessary for correctly calculating the DIC).

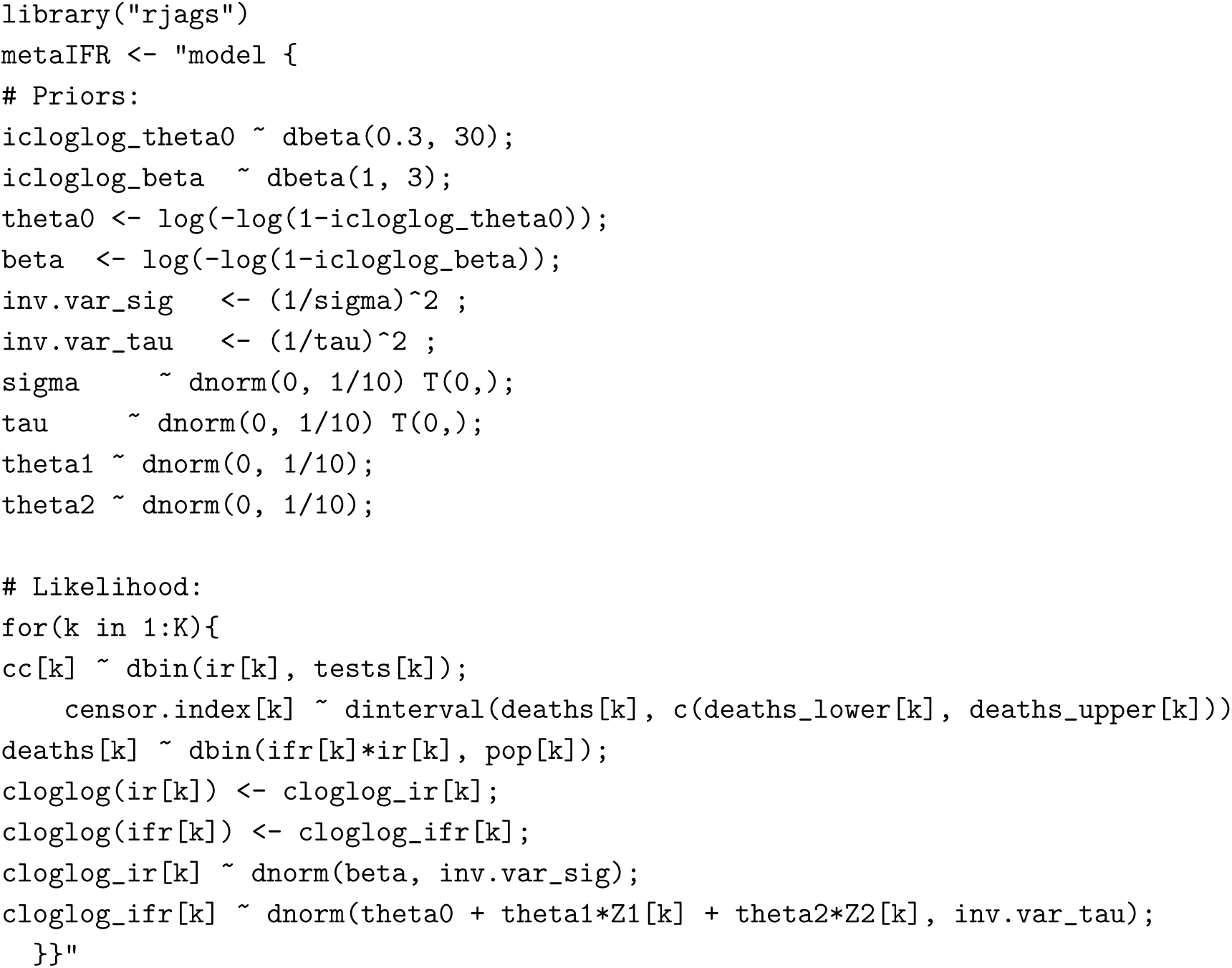

### 6.5 Additional Figures

**Figure 5:**
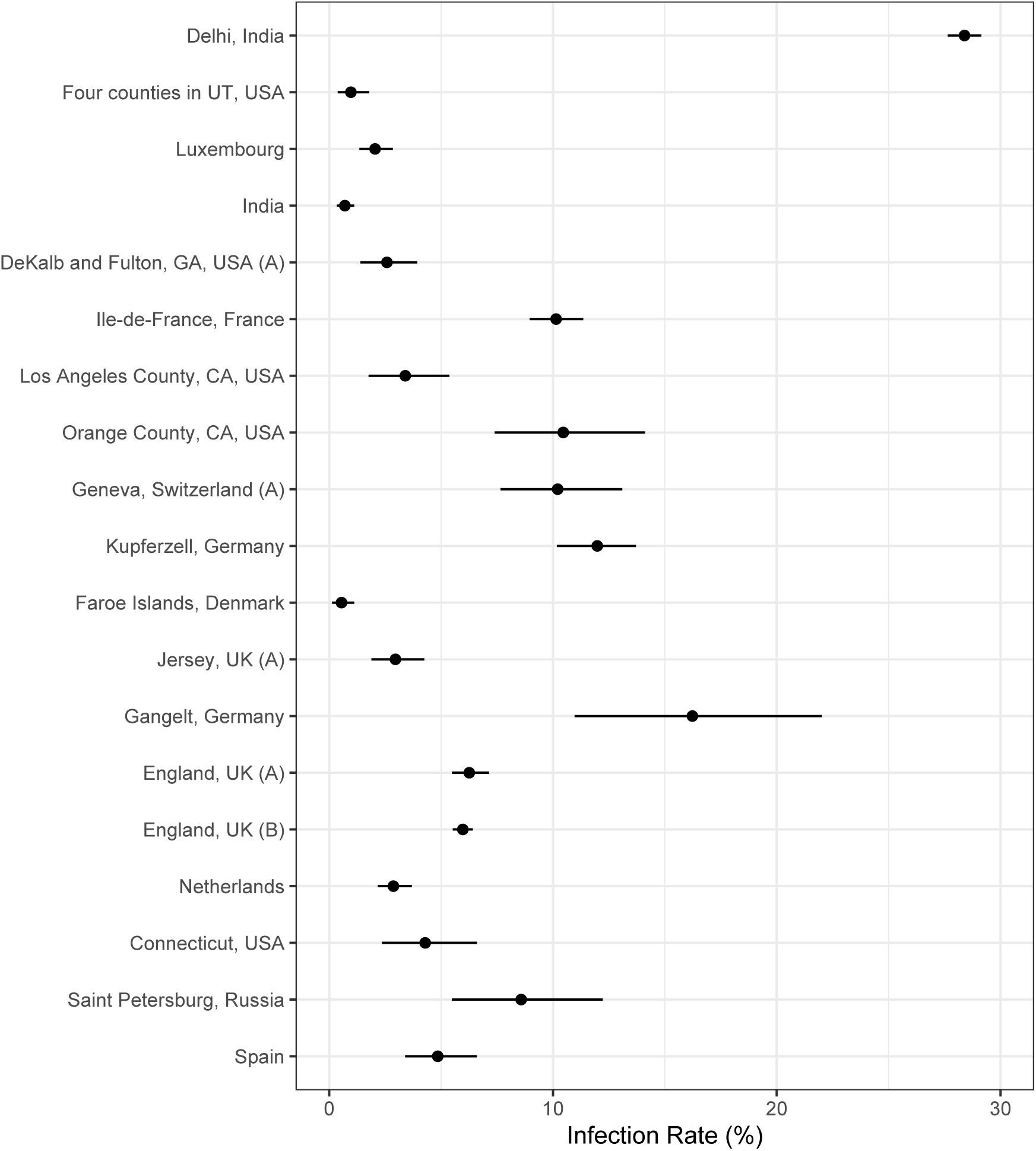
Analysis results from the Chen et al.-based analysis: posterior median estimates for the *IR_k_* variables (for *k* = 1*, . . . ,* 19) with 95% HPD CrIs. Studies are listed from top to bottom in the same order as in Figure 3.

**Figure 6:**
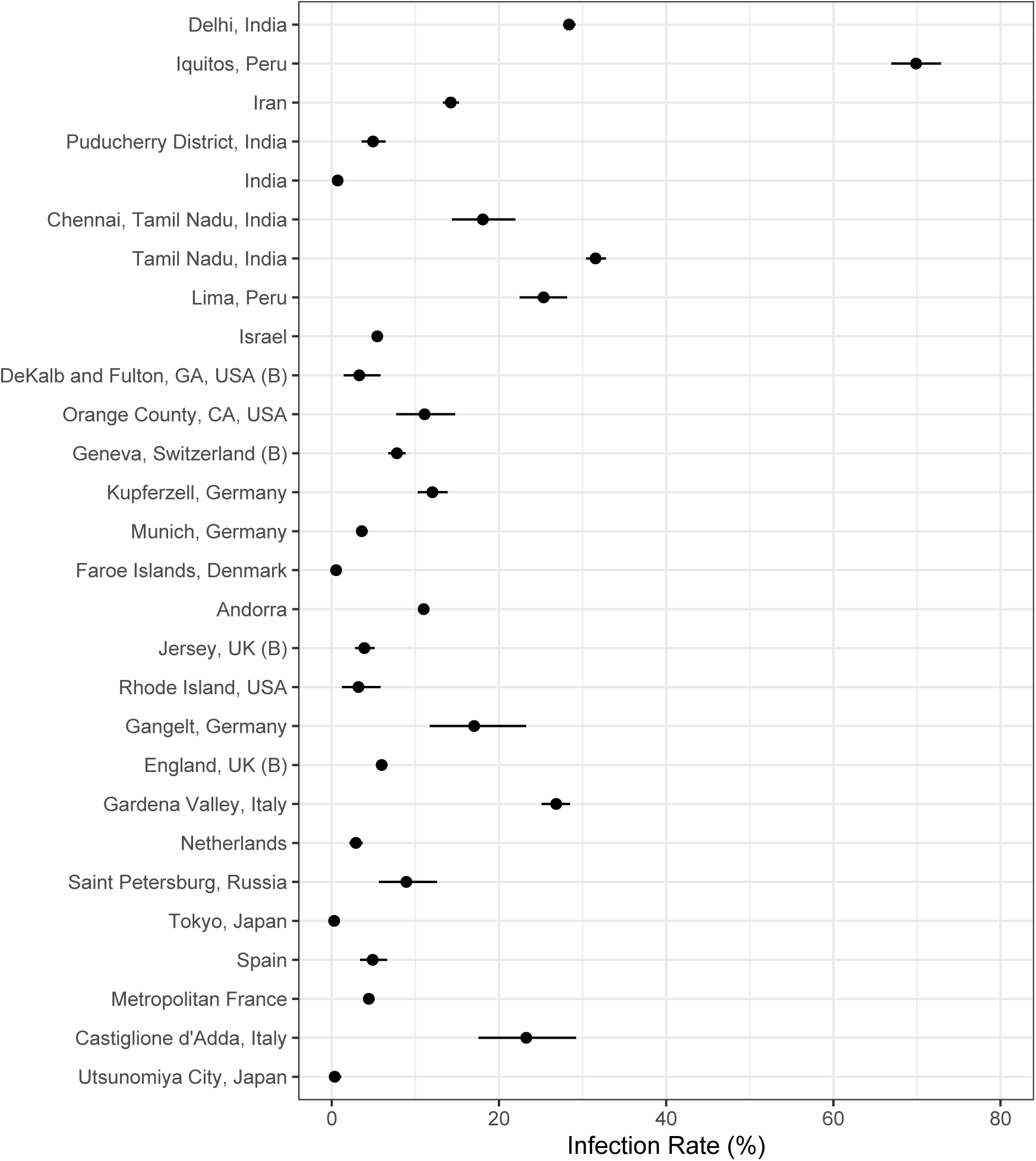
Analysis results from the Serotracker-based analysis: posterior median estimates for the *IR_k_* variables (for *k* = 1*, . . . ,* 28) with 95% HPD CrIs. Studies are listed from top to bottom in the same order as in Figure 4.

### 6.6 Sensitivity analyses

As a sensitivity analysis, we repeated both analyses with an alternative set of priors. For these alternative analyses, we used: g*^−^*^1^(*θ*_0_) *∼ Uniform*(0, 1); g*^−^*^1^(*β*) *∼ Uniform*(0, 1) ; *θ*_1_ *∼ N* (0, 100); *θ*_2_ *∼ N* (0, 100); *σ ∼* half-*N* (0, 100) and *τ ∼* half-*N* (0, 100). The results are plotted in Figures 9 and 10.

We also conducted alternative analyses excluding studies for which the mortality data were not obtained directly from official and reliable sources. See results in Figures 7 and 8. Without the excluded studies, we are unable to provide a reasonable “World” estimate (see the extremely wide credible intervals). However, the “USA” and “EU” estimates are relatively similar.

**Figure 7:**
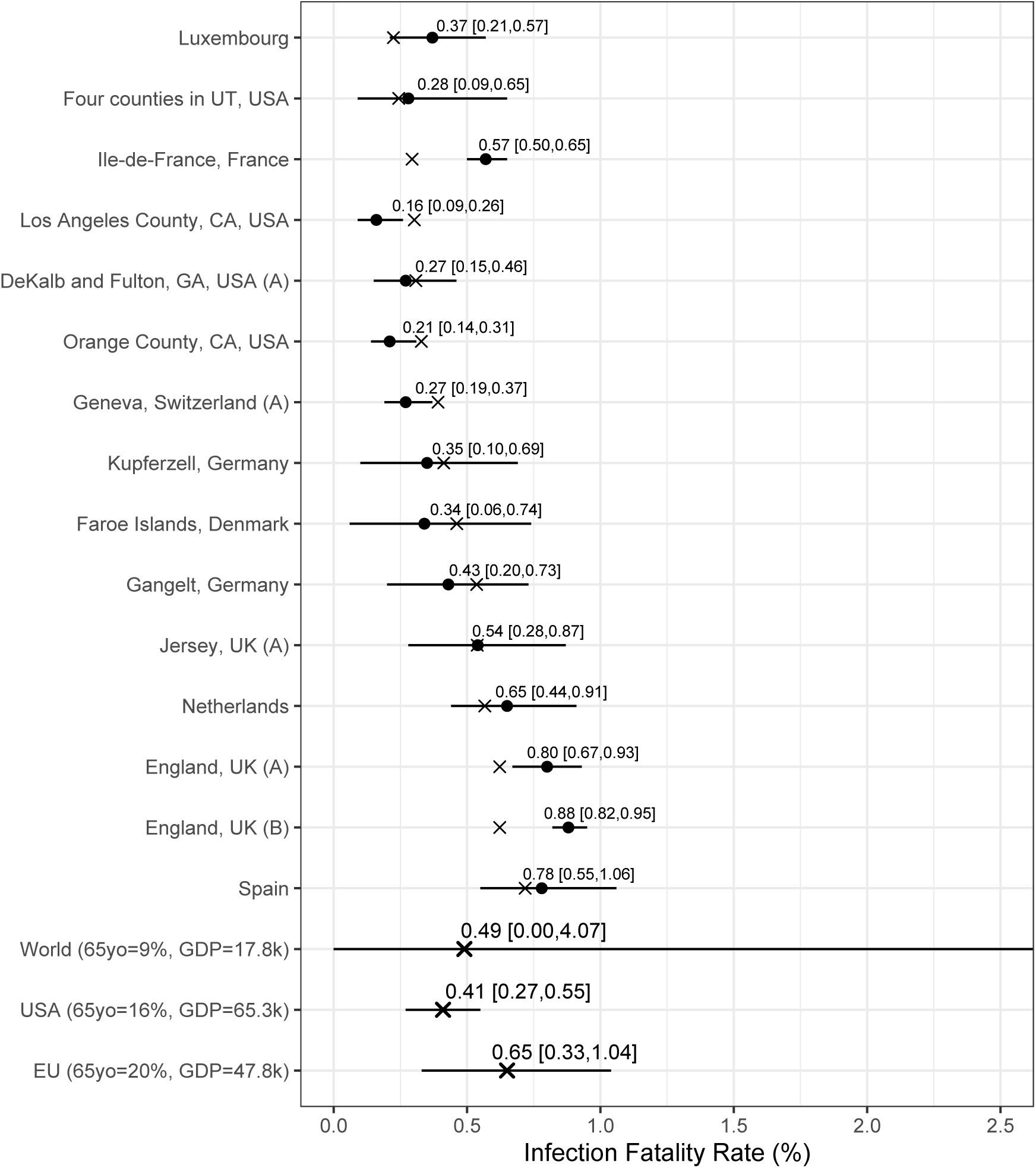
Chen et al.-based analysis results with only those studies for which mortality data was obtained from official sources known to be reliable: posterior median estimates (black circles) for the *IFR_k_* variables (for *k* = 1*, . . . ,* 15) with 95% HPD CrIs. Studies are listed from top to bottom in order of increasing fitted values (these values are indicated by ×). Also plotted, under the labels “World (65 yo=9%, GDP=17.8k)”, “USA (65 yo=16%, GDP=65.3k)”, “EU (65 yo=20%, GDP=47.8k)”, are the posterior median estimate and 95% HPD CrIs for the typical IFR corresponding to values for the proportion of the population aged 65 years and older of 9% and for GDP per capita of $17,811 (the worldwide values), of 16% and of $65,298 (the USA values), and of 20% and of $47,828 (the EU values).

**Figure 8:**
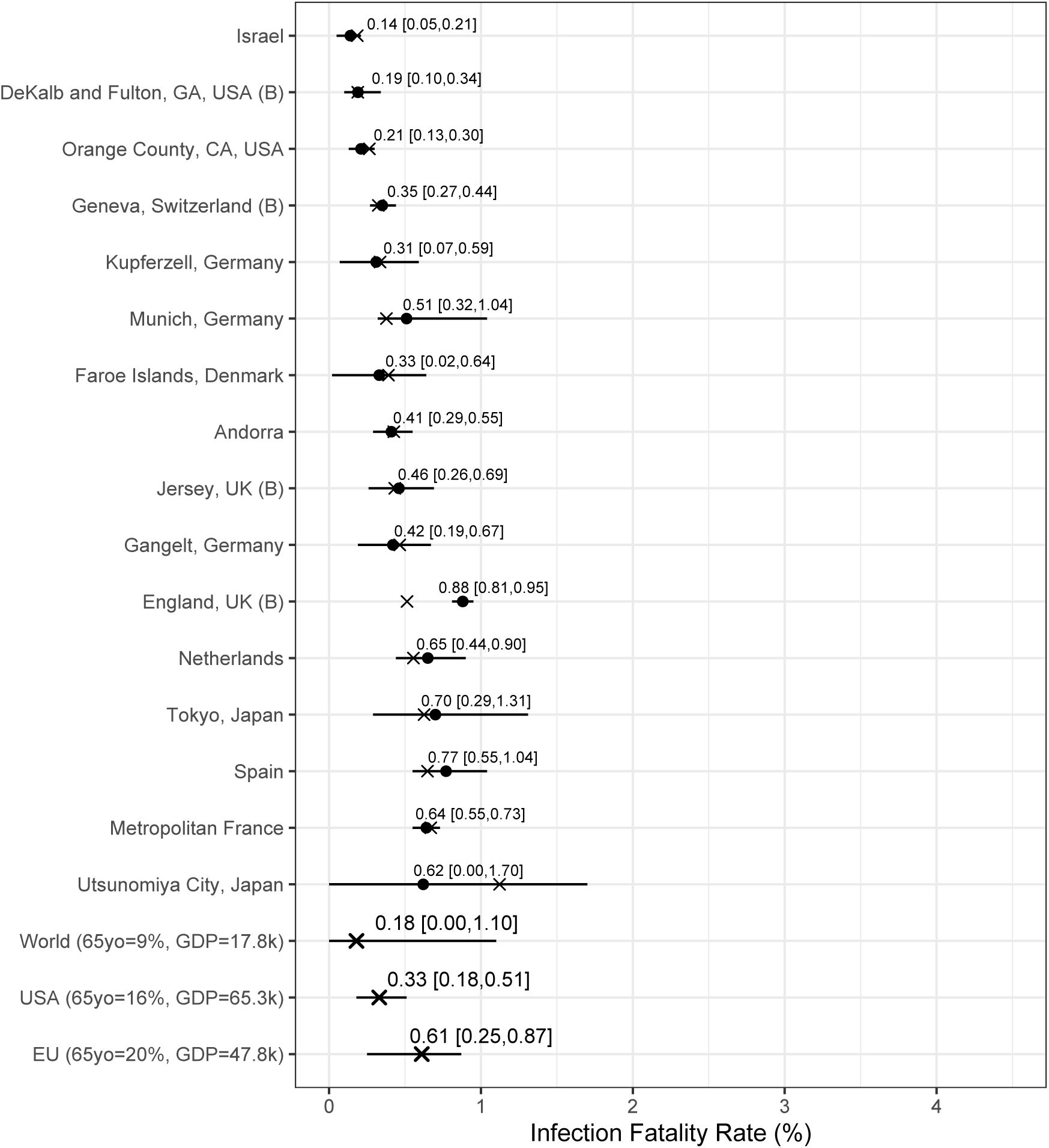
Serotracker-based analysis results with only those studies for which mortality data was obtained from official sources known to be reliable: posterior median estimates (black circles) for the *IFR_k_* variables (for *k* = 1*, . . . ,* 16) with 95% HPD CrIs. Studies are listed from top to bottom in order of increasing fitted values (these values are indicated by ×). Also plotted, under the labels “World (65 yo=9%, GDP=17.8k)”, “USA (65 yo=16%, GDP=65.3k)”, “EU (65 yo=20%, GDP=47.8k)”, are the posterior median estimate and 95% HPD CrIs for the typical IFR corresponding to values for the proportion of the population aged 65 years and older of 9% and for GDP per capita of $17,811 (the worldwide values), of 16% and of $65,298 (the USA values), and of 20% and of $47,828 (the EU values).

**Figure 9:**
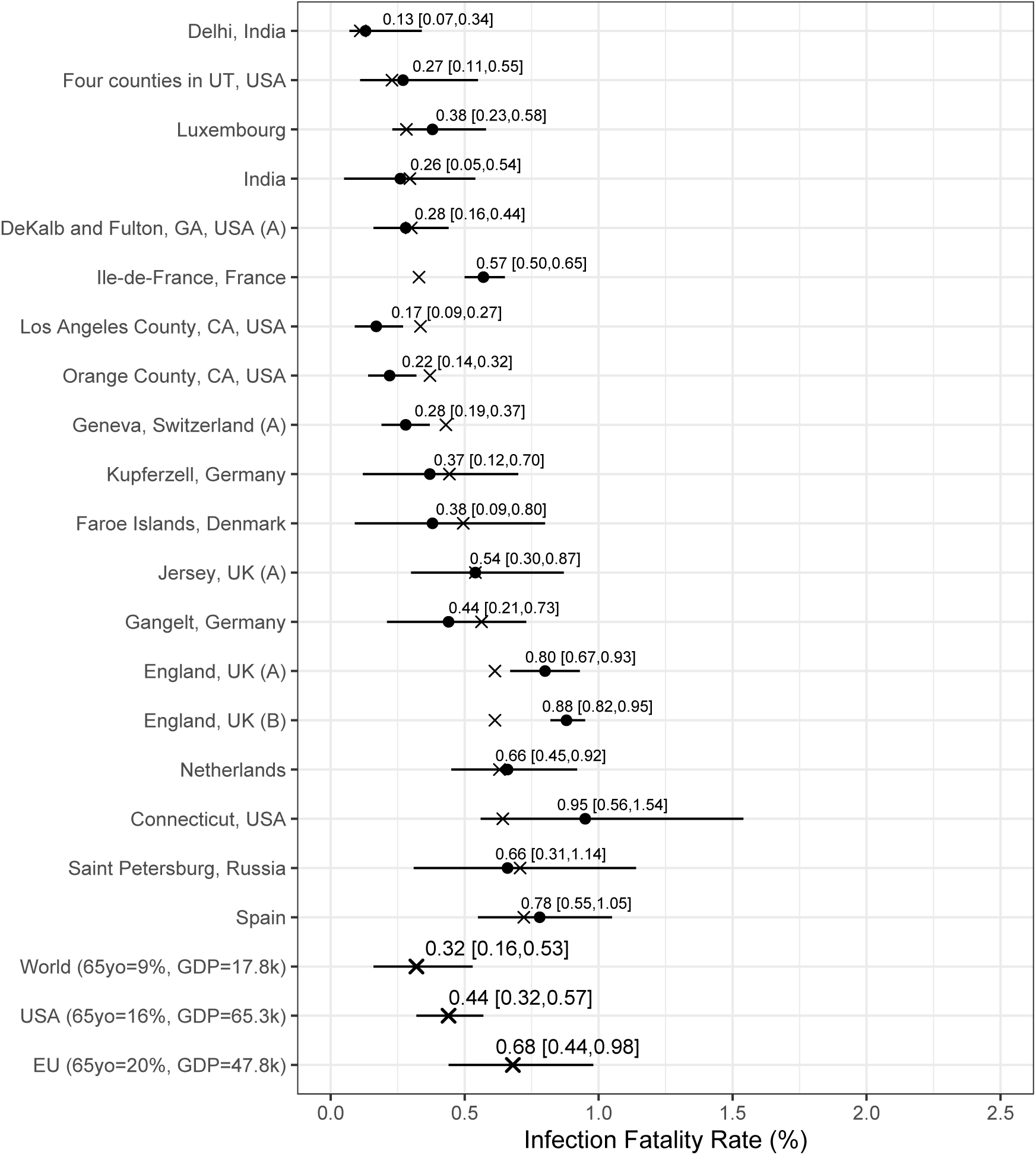
Analysis results from the Chen et al.-based analysis with alternative priors: posterior median estimates (black circles) for the *IFR_k_* variables (for *k* = 1*, . . . ,* 19) with 95% HPD CrIs. Studies are listed from top to bottom according to increasing fitted values (these values are indicated by ). Also plotted, under the labels “World (65 yo=9%, GDP=17.8K)”, “USA (65 yo=16%, GDP=65.3K)”, “EU (65 yo=20%, GDP=47.8K)”, are the posterior median estimate and 95% HPD CrIs for the typical IFR corresponding to values for the proportion of the population aged 65 years and older of 9% and for GDP per capita of $17,811 (the worldwide values), of 16% and of $65,297 (the USA values), and of 20% and of $47,828 (the EU values).

**Figure 10:**
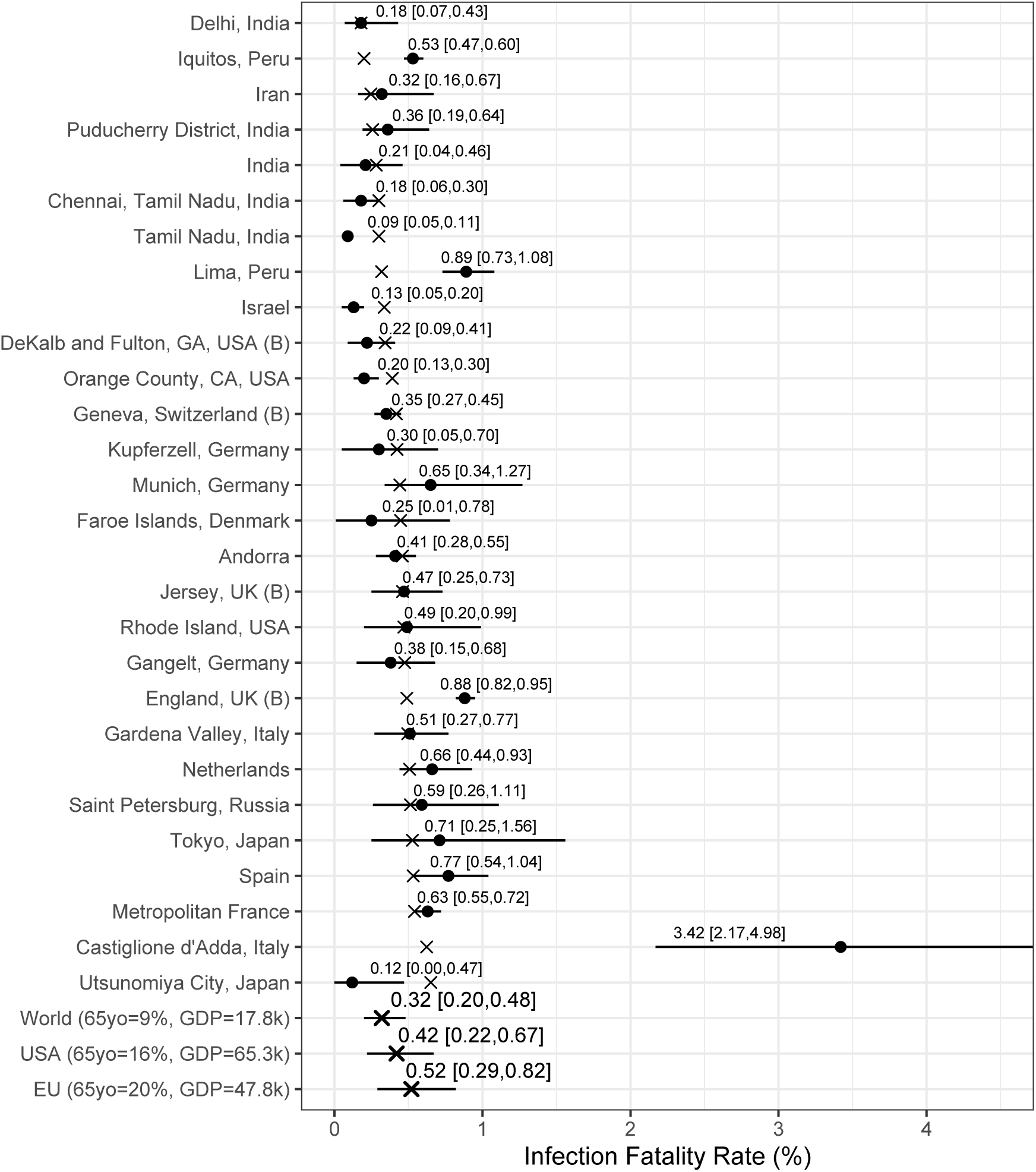
Analysis results from the Serotracker-based analysis with alternative priors: posterior median estimates (black circles) for the *IFR_k_* variables (for *k* = 1*, . . . ,* 28) with 95% HPD CrIs. Studies are listed from top to bottom according to increasing fitted values (these values are indicated by ×). Also plotted, under the labels “World (65 yo=9%, GDP=17.8K)”, “USA (65 yo=16%, GDP=65.3K)”, “EU (65 yo=20%, GDP=47.8K)”, are the posterior median estimate and 95% HPD CrIs for the typical IFR corresponding to values for the proportion of the population aged 65 years and older of 9% and for GDP per capita of $17,811 (the worldwide values), of 16% and of $65,297 (the USA values), and of 20% and of $47,828 (the EU values).

## Footnotes

1 Official regional death numbers for Palestine are available from the Palestinian government dashboard (see https://corona.ps/details; accessed July 28, 2021).

2 Official regional death numbers for Ethiopia have been made available previously (e.g., http://web.archive.org/web/2020*/ https://www.covid19.et/covid-19/ and the Twitter account: https://twitter.com/Harun_Asefa/status/1259069832877793280; accessed August 4, 2021).

3 Malani et al. [76] (“Tamil Nadu”) note that: “Seroprevalence among the elderly (70+: 25.8%) is significantly lower than among the working age populations (age 40-49: 31.6%; *p <* 0.001) or the young (18-29: 30.7%; *p <* 0.001).” Pagani et al. [93] (“Castiglione d’Adda, Italy”) also report that the elderly are more likely to have been infected (“strong association observed between IgG seroprevalence and age”). On the other hand, note that among the 181 participants in the Nawa et al. [89] study (“Utsunomiya City, Japan”) who were aged 65 years or older, none were positive. This suggests that infection in the elderly may be lower than in the general population. However, since the Nawa et al. [89] found only 3 positive cases out of a total of 742 individuals tested, inference on this is limited.

4 For reference, Levin et al. [66] conclude that 87% of the heterogeneity in the IFR (of advanced economies) can be explained by variations in age composition and age-specific prevalence of COVID-19. However, note that the linear regression analysis used to obtain this 87% result is done without an intercept term (see Levin et al. [66] - Figure 6). A linear regression with intercept results in a value of 43%. The intraclass correlation coefficient (ICC) [140] between Levin et al. [66]’s predicted IFRs and the observed IFRs is 0.65 (after removing one outlier, “Portugal”), suggesting that the extent of agreement is reasonably high but nowhere near perfect.

5 While Sullivan et al. [135] used a nationwide representative sampling frame, the results may be subject to sizable selection bias given the low response rate of 12.6%.

6 Bobrovitz et al. [19] conclude that a majority of COVID-19 seroprevalence studies are “at high risk of bias […], often for not statistically correcting for demographics or for test sensitivity and specificity, using non-probability sampling methods, and using non-representative sample frames.” Shook-Sa et al. [123] note that “it is very difficult to estimate the level of bias introduced by convenience sampling.”

